# Causal Bayesian networks to quantify the interactions that influence implementation success

**DOI:** 10.1101/2025.03.04.25323064

**Authors:** Steven Mascaro, Robyn Aitken, Melanie Barwick, Anne B. Chang, Pam Laird, Gloria Lau, Gabrielle B. McCallum, Peter S. Morris, Richard Norman, Maree Toombs, Roz Walker, Andŕe Schultz

## Abstract

Despite the potential of evidence-based medical innovations to improve patient outcomes, their integration remains difficult. Implementation science aims to assist by identifying and deploying effective implementation strategies within complex health care settings. Determinant frameworks, such as the Consolidated Framework for Implementation Research (CFIR), help identify factors influencing implementation success but do not specify mechanisms or methods for selecting optimal strategies. Selection methods are largely empirical, highlighting the need for objective, quantifiable approaches.

We developed causal Bayesian networks (BNs) to model the interdependencies amongst contextual factors, determinants and outcomes with a specific example: the detection and management of chonic wet cough in Indigenous Australian children in primary health care settings. The BNs, informed by CFIR domains and prior qualitative research, quantifies the impact of barriers and enablers on implementation outcomes. The BNs enable predictions of intervention effects, and the assessment and quantification of potential implementation strategies, or a combination of strategies. The BNs are linked to a simple survey that allows implementation strategies to be tailored for each setting and that was administered at several sites across Australia to validate the models. The overall process, including the BNs and surveys, constitutes a generalisable structured workflow for selecting the most promising strategies. We describe the model development and validation, and the broader applicability of our BN-based workflow in implementation science.

## 1 Introduction

Evidence-based medical innovations that could improve the health outcomes of patients are frequently not adopted into routine clinical practice (Institute of Medicine, 2001; Balas and Boren, 2000). Implementation science exists to facilitate the uptake of innovations (like evidence based treatments) in health care institutions using implementation strategies. Health care institutions are by nature complex settings. Therefore, conceptual determinant frameworks like the Consolidated Framework for Implementation Research (CFIR) (Damschroder et al., 2022) have been developed to ensure all factors, contextual and otherwise, that might influence implementation success are considered before a set of implementation strategies are selected or developed for a specific setting. Determinant frameworks provide (or determine) a list of factors that influence implementation outcomes. However, they refrain from specifying the mechanisms by which they do so or providing methods for weighing factors and selecting implementation strategies.

In typical health care settings, a myriad of implementation strategies have the potential to facilitate the uptake of innovations to varying degrees. Selecting the most promising strategies based on what is feasible and likely to effect change can be challenging. Currently, methods for selecting implementation strategies most likely to be impactful exist but are empirical (Perry et al., 2019). There is a clear need to develop objective, quantifiable methods for understanding strategy effectiveness and selecting the most effective implementation strategy (or combination of strategies) in different settings (Ashcraft et al., 2024), particularly in Indigenous settings where there can be great diversity (Loi, 2025). This can potentially be achieved through the study of implementation mechanisms (Vejnoska et al., 2022); specifically by understanding how various contextual factors and determinants in a health care setting influence each other, and then quantifying how various implementation strategies might affect these factors and implementation outcomes.

To this end, we developed a novel approach for applying causal Bayesian networks to quantify and assess implementation strategies and programs. A Bayesian network (BN) is a directed acyclic graph (DAG) that consists of nodes connected by arrows (pointing from ‘parent’ to ‘child’) containing no directed loops (Pearl, 1988; Korb and Nicholson, 2010). Each node represents a random variable with multiple possible states and each node probabilistically depends on its parents. This dependency is quantified by a probability function, typically (and in this paper) a conditional probability table (CPT), specifying the probability distribution of the child given the combined states of all its parents. In a *causal* BN, arrows also represent direct causal influences, allowing them to predict the likely effects of decisions and interventions.^1^ Our approach takes advantage of the key features of causal BNs to provide quantified, mechanistic models capable of providing direct advice to implementers.

To support the development of this approach, and as its first application, we created a causal BN model to map how contextual and other factors might affect the detection and management of chronic wet cough in Indigenous children in primary health care settings. The model quantifies the interaction of individual factors on each other and on implementation outcomes, providing a method for understanding and quantifying the mechanisms through which implementation strategies operate. This allowed us to develop a generalised workflow for creating causal BNs that may be used to support the development of any implementation program.

The proposed model was based on the study of barriers and enablers to the optimal management of chronic wet cough in Indigenous children (Laird et al., 2019), and incorporated all relevant domains of the CFIR (Damschroder et al., 2022). Chronic wet cough is common, particularly in Australian Indigenous children (Laird et al., 2022a), and is often a symptom of lower airway infection that can lead to permanent lung damage (Chang et al., 2018). Optimal management of chronic wet cough leads to improved health outcomes but is often lacking. We have demonstrated that implementing a multi-faceted intervention can facilitate the uptake of evidence-based practice for chronic wet cough and improve health outcomes (Laird et al., 2021). However, studying the barriers and enablers and developing and implementing a multifaceted intervention was a resource-intensive task, even at a single clinic. This highlighted the need for a robust method to select the most effective implementation strategies.

The BN model was validated in primary care clinics in various cultural and geographical regions across Australia as part of a larger project, the Aboriginal and Torres Strait Islander Partnerships for Paediatric Lung Health (APPLE)(Laird et al., 2022b). It is linked to a simple survey that can be administered at primary care clinics to identify and predict implementation outcomes and tailor implementation strategies to each location. In this manuscript, we describe the development and validation of this model as well as the workflow we created for developing such models for implementation endeavours more generally.

## 2 Method

### 2.1 Prior work supporting APPLE BN development

The initial work for designing the implementation program began in 2018 and included qualitative research (including interviews and thematic analysis) to determine the barriers and enablers to the timely detection and guidelines-based-management of chronic wet cough, and is described in detail elsewhere (D’Sylva et al., 2019; Laird et al., 2019). In brief, evidence-based clinical practice guidelines for managing children with chronic wet cough already existed but had not been widely adopted in areas with high incidence. Effective use of the guidelines also required a culturally secure approach in Indigenous settings. Hence, to improve outcomes for affected children, a program was developed and implemented to facilitate the use of clinical practice guidelines in a culturally secure way in primary care settings (Laird et al., 2021).

At the time, our team had no plans to develop a BN nor to analyse the causal mechanisms by which barriers and enablers (or determinants, more generally) influenced implementation outcomes. Hence, the initial identification of determinants was completed without a formal causal modelling process.

The primary outcome measure in the implementation evaluation was children’s respiratory health as measured by parent-proxy Cough-specific Quality of Life (PC-QoL 8-question version)(Newcombe et al., 2013) for children with chronic wet cough. The rationale was that children with chronic wet cough whose symptoms were recognised and correctly managed in primary care would have improved respiratory health 6 weeks to 3 months after being seen in the clinic. An important secondary outcome measure was clinicians’ proficiency in assessing and managing children presenting with respiratory illness. Proficiency was measured through a medical record audit of clinicians recording the presence of cough, cough quality (e.g., wet or dry), cough duration, and correct management (as per guidelines) if the presence of chronic wet cough was detected.

Given Australia’s numerous primary care clinics and their geographical and cultural differences, we adapted the implementation program to each context before testing it in a pseudo-randomised stepped-wedge clinical trial (Laird et al., 2022b). We also explored faster ways to adapt and implement the program, using the CFIR framework (Damschroder et al., 2022) and techniques to analyse the mechanisms behind successful implementations. The latter led directly to using a causal BN to analyse the implementation program.

### 2.2 Development of the APPLE BN

The goal of developing a causal Bayesian Network (BN) was to provide an objective way to identify key factors for improving the detection and management of chronic wet cough in primary care. Initially, the team built a direct representation of the CFIR, but the model was too large and could not easily integrate existing research and thematic analysis. We then shifted to a simpler causal BN focused on specific factors affecting chronic wet cough outcomes (D’Sylva et al., 2019; Laird et al., 2019). These determinants could be divided into two key groups, representing two key pathways to improved management: helping clinicians identify and manage chronic wet cough according to best practice guidelines, and helping families recognise and seek help for chronic wet cough. The causal BN was designed to support clinicians (i.e., the first pathway), as programs had already been designed to support families.

The workflow for developing the APPLE BN is described well by the generalised workflow that we present later in Figure 2 and Section 3.3. One key difference is that the early qualitative research to identify determinants did not at first make use of a determinant framework, as recommended in the workflow. However, the CFIR was later consulted for BN development.

**Figure 1:**
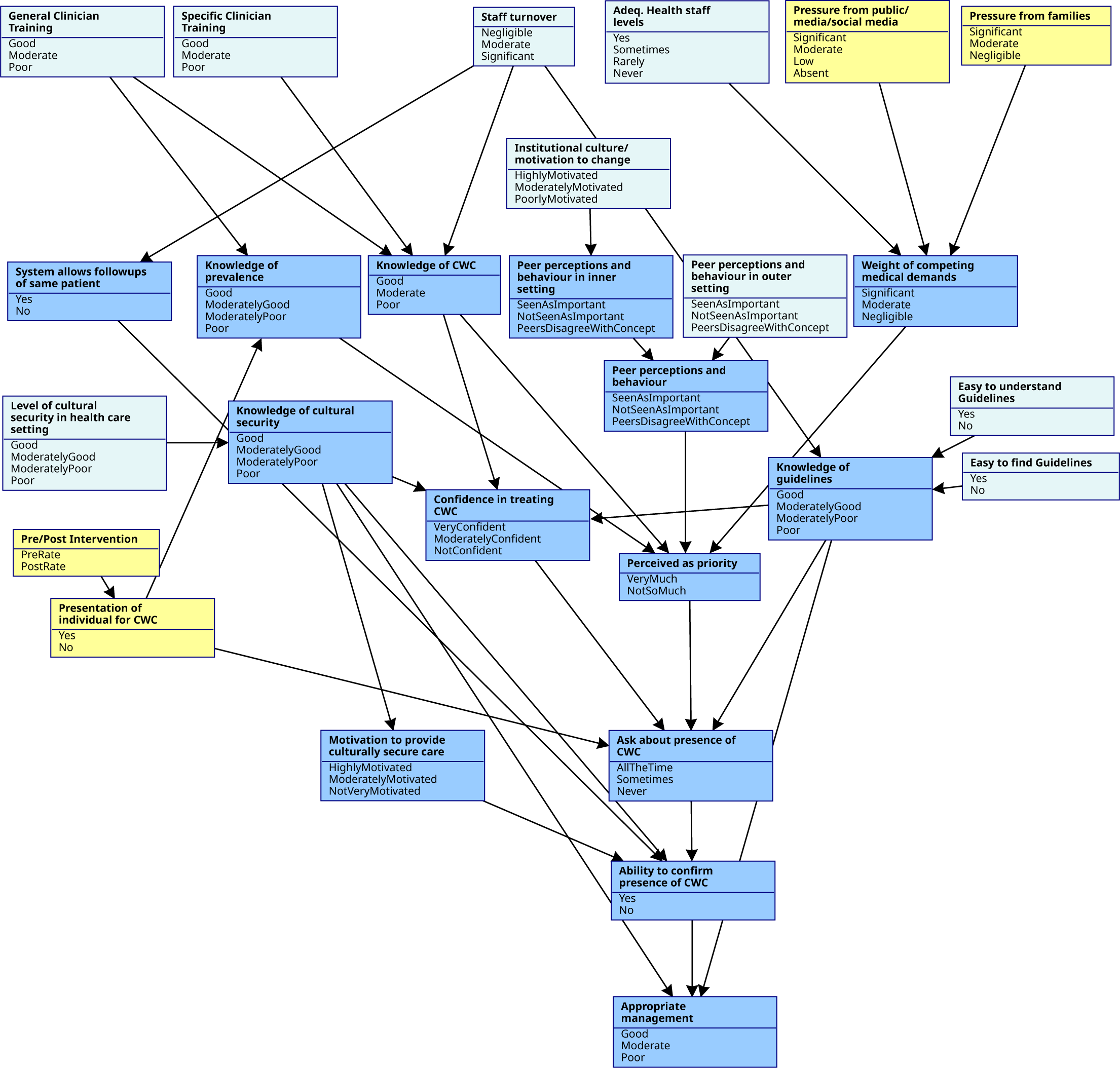
The APPLE BN model illustrates the determinants and causal relationships influencing chronic wet cough management outcomes. The BN structure (nodes and arrows) and states are shown here. The darker blue nodes represent variables with causal parents in the BN, while the lighter blue (root) nodes have none. Yellow nodes highlight important family-related factors not targeted for intervention here, but could be considered in other interventions.

**Figure 2:**
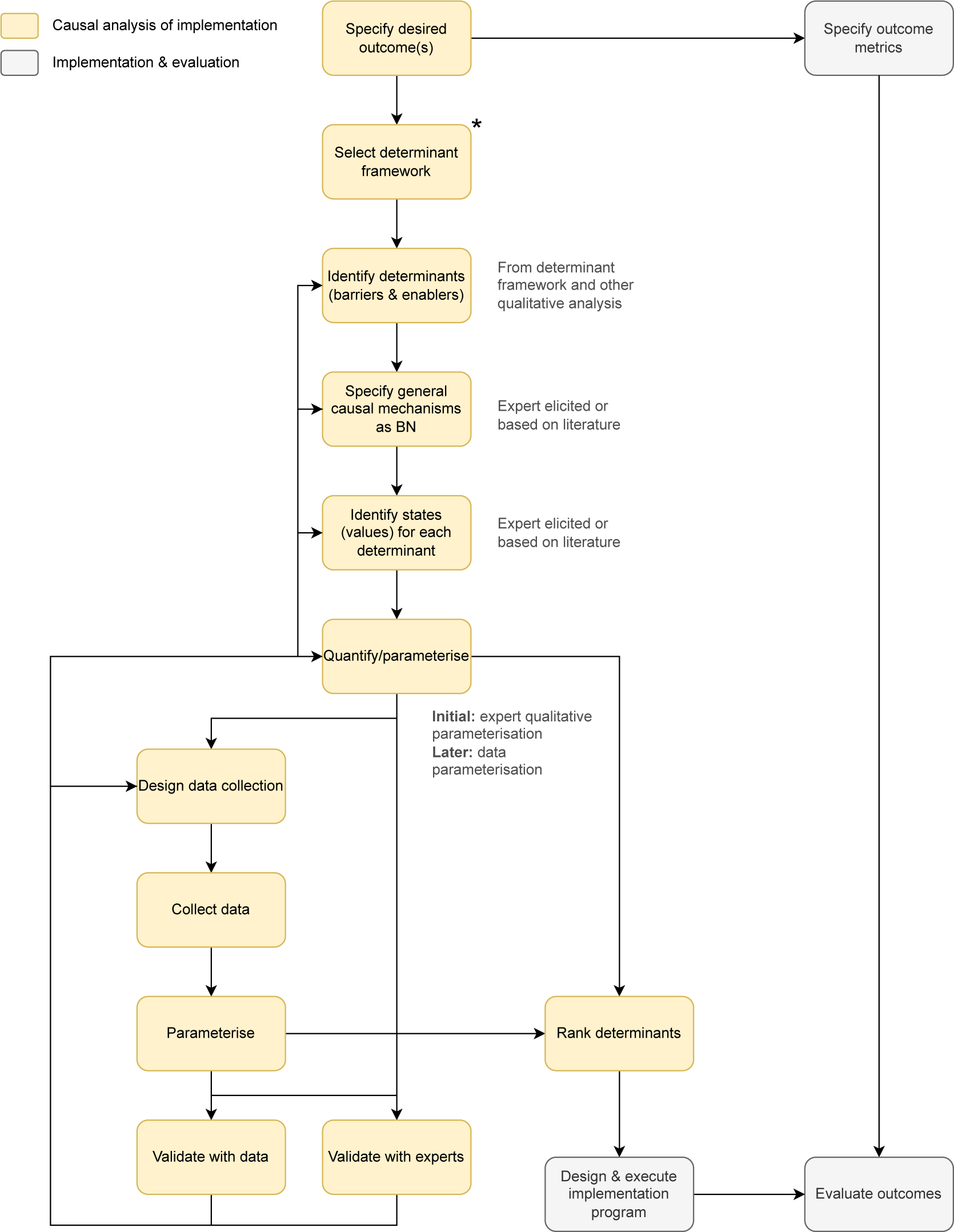
Workflow for developing a causal BN to support an implementation program. Yellow boxes represent steps completed as part of the APPLE BN project. In contrast, grey boxes indicate expected steps in the workflow that were not part of this specific project (though equivalent processes were conducted in other projects). The selection of the determinant framework (marked with an asterisk in the diagram) occurred later in the actual APPLE BN workflow but was ideally positioned earlier, as specified in the diagram.

#### Structure development

The team developed the BN in iterative stages, involving a Bayesian network modeller and three clinicians with chronic wet cough management and implementation science expertise. Two clinicians co-authored a study on barriers and enablers to timely detection and effective management of chronic wet cough in Indigenous children at a single primary care clinic (Laird et al., 2019). The study investigators used semi-structured interviews and focus groups with clinic staff to identify barriers and enablers to timely detection and management of chronic wet cough based on best practice guidelines. The interviews were transcribed and analysed using narrative analysis and thematic grouping. Themes and sub-themes were allocated, discussed and agreed on through an iterative process. NVivo 11 software (QRS International, Melbourne) was used to verify the initial results.

To develop the BN, the modeller first identified key determinants from this work that would serve as the BN’s causal foundations. In a series of workshops focusing on mechanisms, the modeller asked the team to discuss how each determinant affected the primary outcome (i.e., the quality of chronic wet cough management), revising the causal model with the team in real time. In later workshops, the team consulted the CFIR again to identify additional determinants, adding their effects to the model in the same way. Notably, specific strategies and site-specific details were excluded, and a direct link from a determinant to the outcome was avoided if links could be made to mediating pathways that captured its main effects.

#### Structure review

The structure of the causal diagram was reviewed at the beginning of each workshop to ensure the experts had a good understanding of the model and had not missed key determinants or relationships. This involved revising links, identifying new determinants, refining definitions and addressing any inconsistencies. All decisions were made by consensus, guided by the modeller’s advice where required.

#### Initial parameterisation and behaviour

While the causal structure alone offered some guidance on prioritising areas of the implementation program, it can only provide limited insights. Adding parameterisation allows for deeper insights into system behaviour, and allows the impact of determinants to be quantified and ranked. However, obtaining good parameters is difficult due to the qualitative nature of many variables. Although a survey that would help with the parameterisation was planned, parameters were needed prior to this survey — in part, to guide its development. We approached this initial parameterisation in two stages.

In the first stage, the states for each determinant were defined. For each determinant, the modeller asked experts to provide a labelled scale or a set of categories for measuring the determinant. In most cases, these were ordinal scales, such as the scale for the determinant **Ask about presence of chronic wet cough**, which was ‘Never’, ‘Sometimes’ and ‘All the time’. The number of states was limited to four to keep the BN’s conditional probability tables (CPTs) computationally manageable. For example, a CPT with four parents and a child, each with four states, would contain 4^5^ = 1024 parameters.

In the second stage, we used qualitative parameterisation Mascaro et al. (2024a) to capture the model behaviour we expected. This approach specifies parameters that indicate the expected direction of causal influence and the relative magnitude of these changes, without considering numerical accuracy. It can be used when data are absent or sparse to provide a model that captures qualitative causal behaviour, and validates model structures to help guide later data collection efforts. Here, we performed qualitative parameterisation in two phases. In the first phase, the modeller qualitatively parameterised the causal BN to identify any errors or inconsistencies (such as improbable parent state combinations) with the determinants and overall causal structure. In the second phase, experts provided parameter estimates via a Delphi survey. While the CPTs were computationally manageable, they were still too large for direct expert estimation. To simplify this, parent nodes were assumed to influence children independently (often called independence of causal influence, or ICI; Zhang and Yan, 1998) requiring experts to provide only a few parameters per determinant. Specifically, the InterBeta interpolation was used, which involves eliciting 2 distributions, and 1 strength parameter per parent (Mascaro and Woodberry, 2022).

This expert-parameterised BN was used for the structure’s final validation before developing a survey for data collection. The survey was administered at sites participating in the pseudo-randomised trial, where the program was tailored and implemented (Laird et al., 2022b). Survey responses were then used to validate the qualitatively parameterised BN and complete its final parameterisation, as described below.

#### Behaviour and sensitivity review

After parameterising the BN, the experts analyzed its behaviour by testing how the model responded when evidence was applied to the network. Evidence was applied on root nodes (i.e., nodes with no parent influences) to observe their effects propagated through the network to impact the outcome. As before, behaviour was evaluated through a series of expert elicitation workshops.

After these sessions, we examined how sensitive the outcome (i.e., detection and correct management of chronic wet cough) was to each determinant. Determinants were ranked by their influence on the outcome, using mutual information (MI) and causal mutual information (CI). MI comes from information theory, and is the most general measure of association between two variables. In almost all cases here, we provide both the raw MI in bits, and the MI as a percentage of the entropy in the target (which we label ‘MI%’). In this context, MI% measures how much the outcome’s uncertainty is reduced by finding out the state of a determinant on average. In addition, CI% specifies how much the outcome’s uncertainty would be reduced on average if we *intervened* on the determinant (by applying randomisation). We compared this to importance rankings provided by each expert, and compared the experts’ rankings with each other as a measure of expert variability in the assessment of importance.

#### Stakeholder surveys

A survey was developed for staff at primary care clinics to assess the current situation at the clinic where they worked. The survey was based directly on the developed causal BN model, with each question corresponding to one or two determinants in the BN. Most questions were about determinants in isolation rather than about the relationships between determinants. For example, instead of asking, ‘How strong do you believe the relationship to be between Knowledge of CWC and Management of CWC?’, we asked ‘How good is Knowledge of CWC in your clinic?’ and ‘How good is Management of CWC in your clinic?’. This was done for two reasons: 1) it allowed us to *infer* associations from self-reported *observations* of their situations and compare them to our expert estimates, rather than capturing opinions on the causal relationships that might underpin their situations; and 2) asking for opinions on the causal relationships would have unacceptably increased the number of questions and the burden of answering the survey.

The survey was used for two purposes: 1) to determine if the perceptions of local health care staff matched the associations predicted by the model, i.e., if the model captures the causal process correctly; and 2) to provide additional data for the parameterisation of the BN. The survey was hosted online and linked to a QR code, which was sent out to clinic managers for distribution to staff. Staff responding to the survey included doctors, nurses, Indigenous health practitioners, and managers across several locations (see Table 1 for a summary). The survey was composed of two parts. The first part addressed how staff perceived their current situation. The second part described planned elements of the program to be implemented at their site, and asked how they believed the situation might look after such an implementation. By asking for both their current and expected perceptions, deeper insight could be obtained into how strongly they believed determinants were causally related, without asking them about determinant relationships explicitly.

**Table 1:** Count of survey respondents by respondent’s role and location.

| Role | Location |  |  |  | Total |
| --- | --- | --- | --- | --- | --- |
|  | Kutjungka | Halls Creek | Perth | QLD |  |
| Doctor | 2 | 1 | 1 | 2 | 6 |
| Nurse | 3 | 1 | 3 | 3 | 10 |
| AHW |  | 2 |  |  | 2 |
| ALO |  | 1 |  |  | 1 |
| Manager | 1 | 1 |  | 5 | 7 |
| Other |  | 1 |  |  | 1 |
| Total | 6 | 7 | 4 | 10 | 27 |

The survey development process included refining the wording to ensure clarity for respondents, which resulted in revisions to some of the states assigned to the determinants in the model. Responses were collected and processed to align with the model. The associations in the raw responses were checked against the model’s predictions. In addition, a version of the model structure was parameterised using the survey data and compared to the qualitatively parameterised model developed by the expert group. All comparisons used the MI and CI of determinants with the outcome.

#### Parameterisation from survey data

The amount of survey data collected was not sufficient to perform a good quality BN parameterisation using a standard parameter learning approach, such as a count-based (Spiegelhalter and Lauritzen, 1990) or expectation-maximisation-based parameterisation (Dempster et al., 1977). For example, using these techniques with the available data, individual rows in the conditional probability table (CPT) for **Appropriate management** would have an average equivalent sample size of less than one (0.81), and hence would be inadequate. Instead, we trained local Naive Bayes models for each node (i.e., allowing arbitrary but independent relationships with parent nodes), and derived the statistically equivalent CPT. (The average equivalent sample size using this technique would be approximately 9 for each CPT row of the same node.) This is similar in principle to other local structure learning techniques (Friedman and Goldszmidt, 1998), and makes no assumptions about the relationships between parent nodes and the child nodes beyond independence of causal influence (i.e., parent effects are assumed independent).

A question for one node, **Motivation to detect and manage**, was omitted from the survey in error. Fortunately, since the node was internal to the BN and had very simple connections to other nodes (with only one child node), the node could be omitted without harm to the analysis and minimal impact on the BN’s capabilities. Hence, the team chose to proceed without it.

### 2.3 Generalised workflow

After the causal BN was built and refined for the chronic wet cough case study, the team developed the method into a generalised workflow that could potentially be applied to any implementation program in a primary health care setting. The approach involved two steps: first, we explicitly captured the workflow used for APPLE BN in a flowchart; second, we added, clarified or generalised the steps in the flowchart, keeping to the minimum changes needed for the workflow to work well for other similar projects. Consequently, the core of the workflow uses general terms, but directly reflects the workflow used for APPLE BN. The generalised workflow is described in Section 3.3.

## 3 Results

### 3.1 APPLE BN model

The final APPLE BN model structure (comprising its nodes and arrows, with states also visible) is shown in Figure 1. We will refer back to the nodes in this diagram throughout the results, by bolding the variable name when mentioned in the text — for example, **Appropriate management** for the node labelled ‘Appropriate management’ at the bottom of the BN. Aside from minor changes for analysis, the structure and node states are the same as the original version of the APPLE BN that was created via the elicitation process. It captures experts’ final judgements on the most appropriate structure for representing what determinants affect appropriate management and how. The APPLE BN model was parameterised in 3 ways: 1) based on expert estimates alone (the “expert BN”); 2) based on survey data alone (the “survey BN”); and 3) a weighted combination of expert parameters and survey data (the “combined BN” or “final BN”).

The APPLE BN focuses specifically on clinic-based methods of improving chronic wet cough in children, with the BN’s key outcome being appropriate clinical management of chronic wet cough. This appears at the bottom of the model as **Appropriate management**. **Appropriate management** is influenced directly by 3 factors: **Knowledge of cultural security**, **Knowledge of guidelines** and **Ability to confirm presence of CWC**. The first two nodes sit relatively high in the model, directly influenced by determinants identified in the qualitative analysis, including **Level of cultural security in health care setting**, **Easy to understand guidelines**, **Easy to find guidelines** and **Staff turnover**. By contrast, the third direct influencing factor (**Ability to confirm presence of CWC**), sits deep, and is affected by whether the **System allows followups of same patient** and whether the clinician **Asks about presence of CWC**, which in turn is influenced by whether CWC is **Perceived as priority** for the clinician. Asking about presence is also directly affected by **Confidence in treating CWC** and **Knowledge of guidelines**, while perception of priority is influenced by **Knowledge of CWC** and its prevalence, **Peer perceptions and behaviour** in the outer and inner settings and **Weight of competing medical demands**.

The nodes just described (darker blue in the figure) are all consequences of other causes further up the causal pathways. Some of these are determinants identified during the qualitative analysis(Laird et al., 2019), while the remainder are key mediating nodes conceived and included during the elicitation process. The causes at the top of the graph (i.e., the root nodes in lighter blue) are all determinants from the qualitative analysis. While it may be possible to intervene on any node in the BN, the root nodes will often be the most directly modifiable by any practical intervention.

#### 3.1.1 Model Behaviour

We will begin by analysing the patterns in the individual conditional probability tables (CPTs) of the combined APPLE BN, which will help us understand the local relationships in the model (i.e., how each determinant is affected by its immediate causes). We will also compare the CPTs of the combined BN with those of the expert and survey BNs. While the expert BN’s CPTs will be constrained to assumed linear relationships, the data-informed CPTs from the survey and combined BNs may reveal more complex non-linear patterns. Even still, there will be no interactions between parents due to the Näıve Bayes-based training. Following this, we will examine the BN’s high-level, global behaviour under different scenarios and interventions.

The behaviour of the final node, **Appropriate management**, is of greatest interest and its CPT contributes most to how this outcome responds to other nodes in the BN. This CPT is shown in Table 2 for the combined BN, and, for comparison, is followed by the corresponding CPTs from the expert and survey BNs. The heatmap shows the clear influence of the **Ability to confirm presence of CWC** for all three BNs. In particular, the top half of the table (where the variable has the value *Yes*) differs from the bottom half (where it has the value *No*). The influence of **Knowledge of guidelines** is more minor in every CPT (there is not a great difference between every group of 4 rows). Meanwhile, **Knowledge of cultural security** has a much stronger influence in the survey BN than in the expert BN, with the combined BN, of course, being a combination of these two strengths of influence (compare rows 1 and 2, with rows 3 and 4, for example, and so on within every block of 4 rows).

**Table 2:**
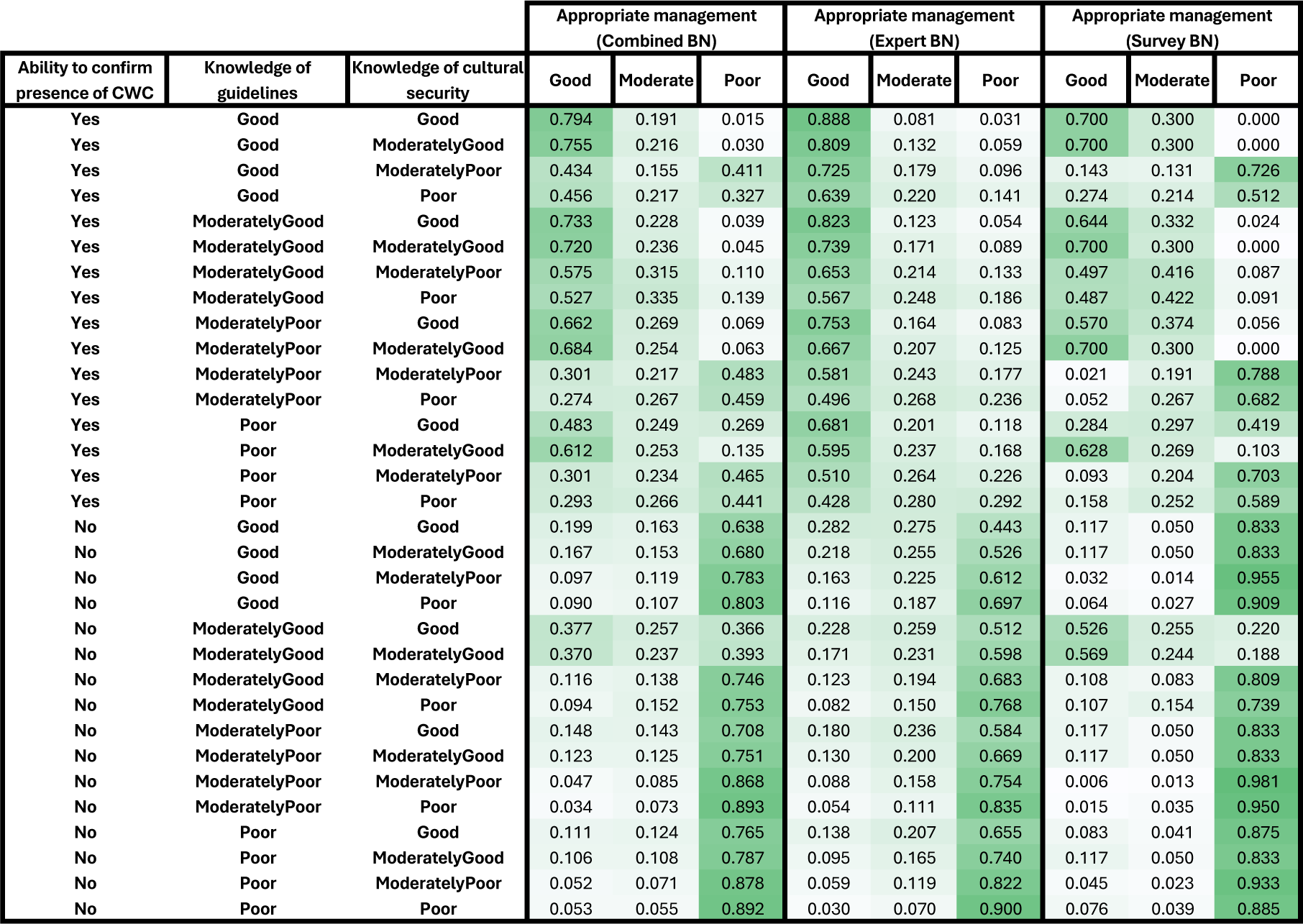
The CPT for **Appropriate management** that appears in the Combined BN (columns 4-6), Expert BN (columns 7-9) and Survey BN (columns 10-12). Each row represents a possible combination of the parent determinant states (labelled in columns 1-3). Higher probabilities are shaded a darker green.

| Ability to confirm presence of CWC | Knowledge of guidelines | Knowledge of cultural security | Appropriate management (Combined BN) |  |  | Appropriate management (Expert BN) |  |  | Appropriate management (Survey BN) |  |  |
| --- | --- | --- | --- | --- | --- | --- | --- | --- | --- | --- | --- |
|  |  |  | Good | Moderate | Poor | Good | Moderate | Poor | Good | Moderate | Poor |
| Yes | Good | Good | 0.794 | 0.191 | 0.015 | 0.888 | 0.081 | 0.031 | 0.700 | 0.300 | 0.000 |
| Yes | Good | ModeratelyGood | 0.755 | 0.216 | 0.030 | 0.809 | 0.132 | 0.059 | 0.700 | 0.300 | 0.000 |
| Yes | Good | ModeratelyPoor | 0.434 | 0.155 | 0.411 | 0.725 | 0.179 | 0.096 | 0.143 | 0.131 | 0.726 |
| Yes | Good | Poor | 0.456 | 0.217 | 0.327 | 0.639 | 0.220 | 0.141 | 0.274 | 0.214 | 0.512 |
| Yes | ModeratelyGood | Good | 0.733 | 0.228 | 0.039 | 0.823 | 0.123 | 0.054 | 0.644 | 0.332 | 0.024 |
| Yes | ModeratelyGood | ModeratelyGood | 0.720 | 0.236 | 0.045 | 0.739 | 0.171 | 0.089 | 0.700 | 0.300 | 0.000 |
| Yes | ModeratelyGood | ModeratelyPoor | 0.575 | 0.315 | 0.110 | 0.653 | 0.214 | 0.133 | 0.497 | 0.416 | 0.087 |
| Yes | ModeratelyGood | Poor | 0.527 | 0.335 | 0.139 | 0.567 | 0.248 | 0.186 | 0.487 | 0.422 | 0.091 |
| Yes | ModeratelyPoor | Good | 0.662 | 0.269 | 0.069 | 0.753 | 0.164 | 0.083 | 0.570 | 0.374 | 0.056 |
| Yes | ModeratelyPoor | ModeratelyGood | 0.684 | 0.254 | 0.063 | 0.667 | 0.207 | 0.125 | 0.700 | 0.300 | 0.000 |
| Yes | ModeratelyPoor | ModeratelyPoor | 0.301 | 0.217 | 0.483 | 0.581 | 0.243 | 0.177 | 0.021 | 0.191 | 0.788 |
| Yes | ModeratelyPoor | Poor | 0.274 | 0.267 | 0.459 | 0.496 | 0.268 | 0.236 | 0.052 | 0.267 | 0.682 |
| Yes | Poor | Good | 0.483 | 0.249 | 0.269 | 0.681 | 0.201 | 0.118 | 0.284 | 0.297 | 0.419 |
| Yes | Poor | ModeratelyGood | 0.612 | 0.253 | 0.135 | 0.595 | 0.237 | 0.168 | 0.628 | 0.269 | 0.103 |
| Yes | Poor | ModeratelyPoor | 0.301 | 0.234 | 0.465 | 0.510 | 0.264 | 0.226 | 0.093 | 0.204 | 0.703 |
| Yes | Poor | Poor | 0.293 | 0.266 | 0.441 | 0.428 | 0.280 | 0.292 | 0.158 | 0.252 | 0.589 |
| No | Good | Good | 0.199 | 0.163 | 0.638 | 0.282 | 0.275 | 0.443 | 0.117 | 0.050 | 0.833 |
| No | Good | ModeratelyGood | 0.167 | 0.153 | 0.680 | 0.218 | 0.255 | 0.526 | 0.117 | 0.050 | 0.833 |
| No | Good | ModeratelyPoor | 0.097 | 0.119 | 0.783 | 0.163 | 0.225 | 0.612 | 0.032 | 0.014 | 0.955 |
| No | Good | Poor | 0.090 | 0.107 | 0.803 | 0.116 | 0.187 | 0.697 | 0.064 | 0.027 | 0.909 |
| No | ModeratelyGood | Good | 0.377 | 0.257 | 0.366 | 0.228 | 0.259 | 0.512 | 0.526 | 0.255 | 0.220 |
| No | ModeratelyGood | ModeratelyGood | 0.370 | 0.237 | 0.393 | 0.171 | 0.231 | 0.598 | 0.569 | 0.244 | 0.188 |
| No | ModeratelyGood | ModeratelyPoor | 0.116 | 0.138 | 0.746 | 0.123 | 0.194 | 0.683 | 0.108 | 0.083 | 0.809 |
| No | ModeratelyGood | Poor | 0.094 | 0.152 | 0.753 | 0.082 | 0.150 | 0.768 | 0.107 | 0.154 | 0.739 |
| No | ModeratelyPoor | Good | 0.148 | 0.143 | 0.708 | 0.180 | 0.236 | 0.584 | 0.117 | 0.050 | 0.833 |
| No | ModeratelyPoor | ModeratelyGood | 0.123 | 0.125 | 0.751 | 0.130 | 0.200 | 0.669 | 0.117 | 0.050 | 0.833 |
| No | ModeratelyPoor | ModeratelyPoor | 0.047 | 0.085 | 0.868 | 0.088 | 0.158 | 0.754 | 0.006 | 0.013 | 0.981 |
| No | ModeratelyPoor | Poor | 0.034 | 0.073 | 0.893 | 0.054 | 0.111 | 0.835 | 0.015 | 0.035 | 0.950 |
| No | Poor | Good | 0.111 | 0.124 | 0.765 | 0.138 | 0.207 | 0.655 | 0.083 | 0.041 | 0.875 |
| No | Poor | ModeratelyGood | 0.106 | 0.108 | 0.787 | 0.095 | 0.165 | 0.740 | 0.117 | 0.050 | 0.833 |
| No | Poor | ModeratelyPoor | 0.052 | 0.071 | 0.878 | 0.059 | 0.119 | 0.822 | 0.045 | 0.023 | 0.933 |
| No | Poor | Poor | 0.053 | 0.055 | 0.892 | 0.030 | 0.070 | 0.900 | 0.076 | 0.039 | 0.885 |

The optimal scenario in our CPT is represented in the top row, where each parent node is in its best possible state. This combination of parent states leads to a 79% chance of good management and a 1.5% chance of poor management in the combined BN. This best possible outcome still allows some room for improvement (and is lower than in the expert BN’s CPT) but is still relatively high. According to the CPT, the worst prospect is a 5.3% chance of appropriate management and an 89.2% chance of poor management. Hence, a large amount of variation is possible according to this CPT alone. This large variation in the best and worst cases is also clearly visible in both the expert and survey BN CPTs. The key difference in the survey BN is the stronger impact of **Knowledge of cultural security**.

We can also compute the average distribution of the outcome conditional on each parent node in isolation.^2^ These averaged tables are shown in Table 3, along with the estimated strength of influence (equal to the MI between the selected parent and the outcome). These results confirm our earlier statements about each parent’s relative strength. In particular, **Ability to confirm presence of CWC** has a clear influence, **Knowledge of cultural security** has a strong but lesser influence, while the influence of **Knowledge of guidelines** is mixed, though not absent.

**Table 3:**
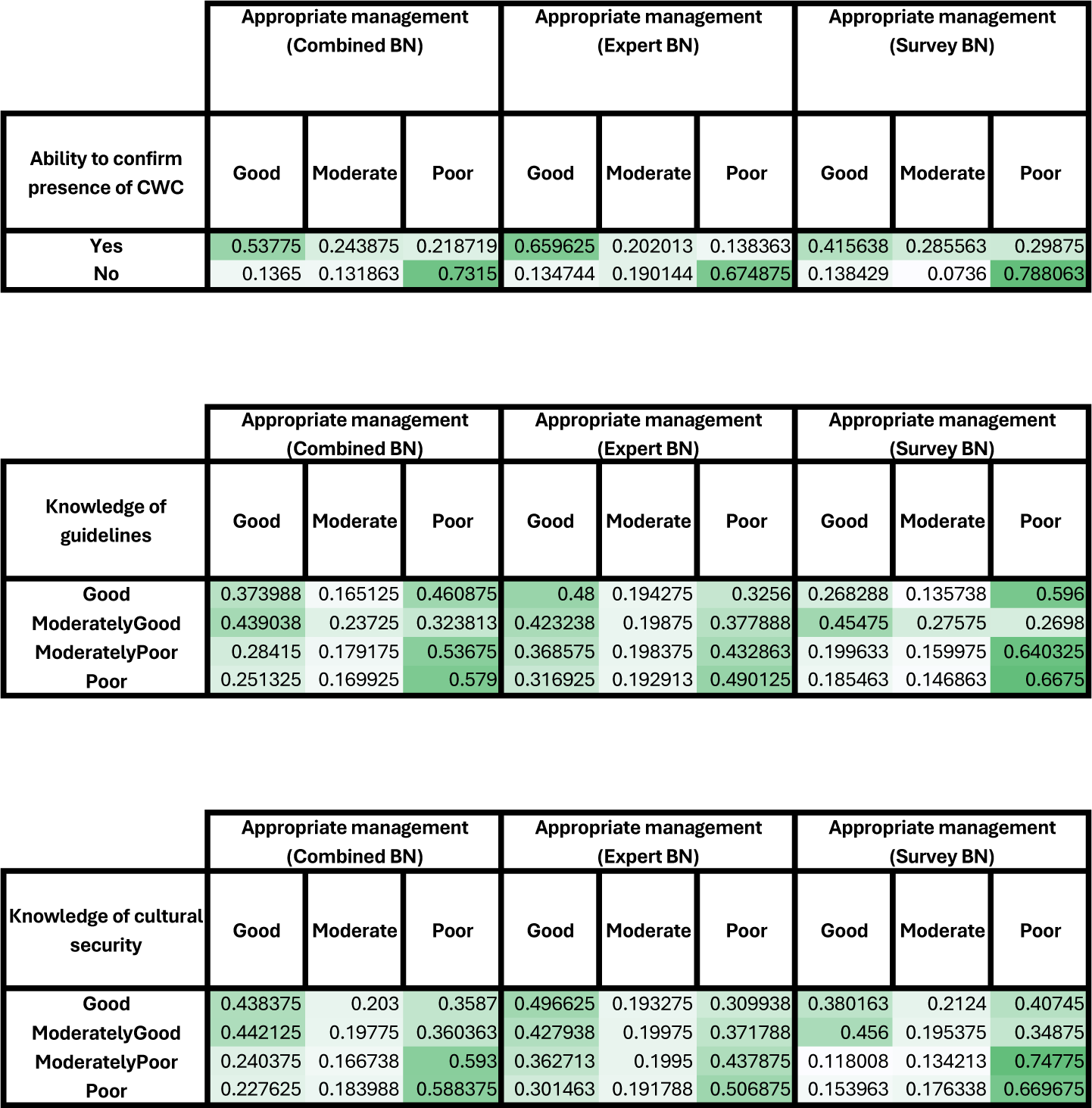
The average distributions for **Appropriate management** conditioned on each parent node separately. For example, the first row in the top table represents the average of all rows from Table 2 where **Ability to confirm presence of CWC** is set to **Yes**; similarly, the second row is the average for all rows where it is set to **No**. This approach isolates the influence of each parent node by averaging probabilities across all other states, providing a clearer view of how each determinant impacts the outcome individually.

|  | Appropriate management<br>(Combined BN) |  |  | Appropriate management<br>(Expert BN) |  |  | Appropriate management<br>(Survey BN) |  |  |
| --- | --- | --- | --- | --- | --- | --- | --- | --- | --- |
| Ability to confirm<br>presence of CWC | Good | Moderate | Poor | Good | Moderate | Poor | Good | Moderate | Poor |
| Yes | 0.53775 | 0.243875 | 0.218719 | 0.659625 | 0.202013 | 0.138363 | 0.415638 | 0.285563 | 0.29875 |
| No | 0.1365 | 0.131863 | 0.7315 | 0.134744 | 0.190144 | 0.674875 | 0.138429 | 0.0736 | 0.788063 |
| Knowledge of<br>guidelines | Good | Moderate | Poor | Good | Moderate | Poor | Good | Moderate | Poor |
| Good | 0.373988 | 0.165125 | 0.460875 | 0.48 | 0.194275 | 0.3256 | 0.268288 | 0.135738 | 0.596 |
| ModeratelyGood | 0.439038 | 0.23725 | 0.323813 | 0.423238 | 0.19875 | 0.377888 | 0.45475 | 0.27575 | 0.2698 |
| ModeratelyPoor | 0.28415 | 0.179175 | 0.53675 | 0.368575 | 0.198375 | 0.432863 | 0.199633 | 0.159975 | 0.640325 |
| Poor | 0.251325 | 0.169925 | 0.579 | 0.316925 | 0.192913 | 0.490125 | 0.185463 | 0.146863 | 0.6675 |
| Knowledge of cultural<br>security | Good | Moderate | Poor | Good | Moderate | Poor | Good | Moderate | Poor |
| Good | 0.438375 | 0.203 | 0.3587 | 0.496625 | 0.193275 | 0.309938 | 0.380163 | 0.2124 | 0.40745 |
| ModeratelyGood | 0.442125 | 0.19775 | 0.360363 | 0.427938 | 0.19975 | 0.371788 | 0.456 | 0.195375 | 0.34875 |
| ModeratelyPoor | 0.240375 | 0.166738 | 0.593 | 0.362713 | 0.1995 | 0.437875 | 0.118008 | 0.134213 | 0.74775 |
| Poor | 0.227625 | 0.183988 | 0.588375 | 0.301463 | 0.191788 | 0.506875 | 0.153963 | 0.176338 | 0.669675 |

The strength of influence of each parent based on the average effect of the parent in the child’s CPT is shown in Figure 4 in Appendix Appendix A. The thickness of each arrow corresponds to the strength of the parent’s influence. We can see a generally thick set of arrows between **Level of cultural security in health care setting** and **Appropriate management**, suggesting this is likely an important path. Arrows on the right of the graph are generally weaker, including arrows that enter and exit **Weight of competing medical demands**, suggesting these may not play a key role. Similarly, the link between **Perceived as priority** and **Ask about presence of CWC** is not as strong as expected.

**Figure 3:**
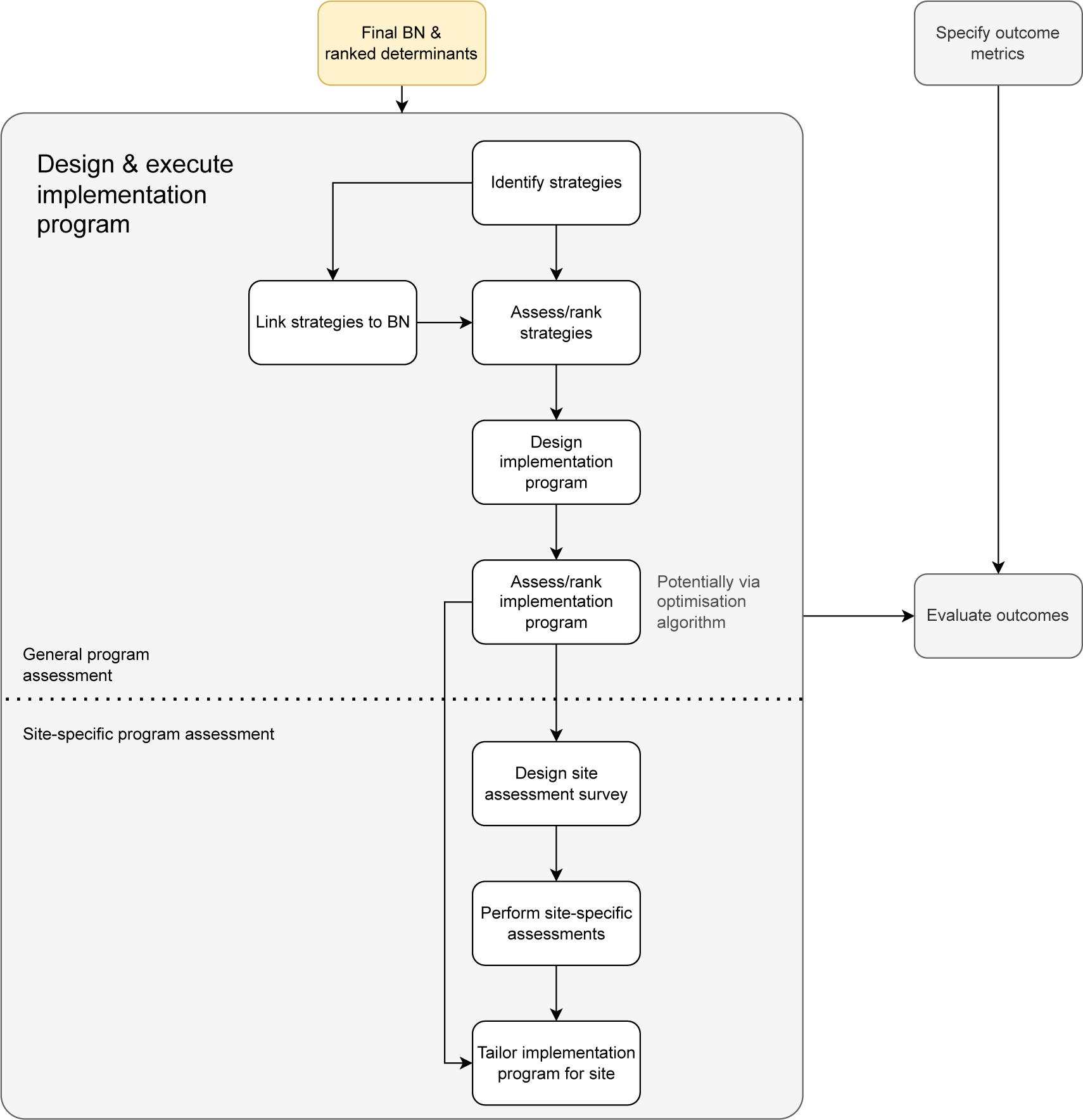
Potential future stage of the generalised workflow for implementation design. This diagram represents a potential future stage in the generalized workflow, focusing on the design of the implementation approach. It builds upon the final yellow box in Figure 2 and elaborates on the *Design & execute implementation program* step. The workflow is divided into two parts: the first half addresses general program assessment, which is not site-specific, while the second half focuses on designing site-specific implementation programs. Typically, the site-specific design phase would follow the general program assessment from the first half, providing a structured path for refining and tailoring implementation strategies.

Finally, we look at the broader impact of determinants on **Appropriate management** in the combined BN. We compare 3 scenarios: the average case (i.e., marginal probabilities only), the best case for all root nodes (i.e., with each root node set to the state that maximises the chance of good **Appropriate management**) and the worst case for all root nodes. The distributions over **Appropriate management** are shown for each case in Table 4. The variation here must strictly be smaller than the variation we saw in the CPT, as the influence of all root nodes can only operate indirectly through **Appropriate management**’s parents. The range is indeed more restricted, with the probabilities of good **Appropriate management** spanning from 19% to 60% (a range of 41%), with a similar variation for poor management. This compares to the wider 5% to 79% interval (a range of 74%) for the CPT (see top and bottom rows of column 4 in Table 2). The key limitation is **Ability to confirm presence of CWC**, which reaches a maximum probability of 75% in the best scenario. If this parent node is manually set to its best state, the chance of good appropriate management in the best scenario would increase to 73%, highlighting its significant influence on the outcome.

**Table 4:** The probability distribution for **Appropriate Management** under three scenarios for all root nodes: 1) best case, in which each root node is set to its best state; 2) average case, in which all root nodes remain unset, and use their prior distributions; and 3) worst case, in which each root node is set to its worst state. This comparison highlights how different root node configurations impact the likelihood of achieving appropriate management, providing a sense of the model’s sensitivity to the root nodes.

|  | Appropriate management<br>(Combined BN) |  |  |
| --- | --- | --- | --- |
|  | Good | Moderate | Poor |
| Best Case | 0.6 | 0.21 | 0.19 |
| Avg Case | 0.39 | 0.19 | 0.42 |
| Worst Case | 0.19 | 0.15 | 0.66 |

### 3.2 Evaluation of the model

This evaluation determines whether the model accurately reflects how primary care clinics respond to potential interventions or programs. To achieve this, we assessed the following aspects of the model:

1. How well the model represents the causal influence of determinants on the appropriate management of chronic wet cough
2. Whether the causal arrows in the model are supported by evidence, and whether there are missing causal arrows
3. Whether the model captures the key determinants
4. Whether we have used appropriate definitions and possible states for the nodes

A qualitative validation approach was used to evaluate the model. This process began before the development of the APPLE BN, with the determinants identified during the initial qualitative analysis being validated for completeness at that stage (Laird et al., 2019). The BN-specific survey provided further validation by exploring facilitators or barriers to implementation that earlier questions may have missed. Since the survey questions correspond precisely to the determinants in the expert BN, this provides a qualitative validation of the coverage. The responses are summarised in Table 5. Most suggestions (in black) were already covered well by existing determinants within the BN, often identical to an existing variable or suitable substitutes for one or more variables. Several determinants (in purple) were not in the BN but were out of scope; these primarily related to the patient rather than the health practitioner. A few suggested factors (in red) were in scope and were not direct substitutes for existing determinants in the model. All such factors influenced existing determinants in the BN, and, perhaps of particular interest, the determinants they influenced were either root nodes or their direct children. This suggests that no major pathway to the final node has been overlooked.

**Table 5:** Additional suggested facilitators & barriers. This table categorises suggestions from the original free text, coded to align with the BN variable names. **Black:** Suggestions matched one or more variables in the BN. **Purple:** Suggestions considered out of scope for the BN. **Red:** Suggestions did not directly match BN variables, but were related to existing variables (often as causes of those existing variables).

| Suggested facilitators | Suggested barriers |
| --- | --- |
| None | Weight of competing medical demands |
| Institutional culture/motivation to change | None |
| None | None |
| **Culturally appropriate educational resources** (closest: Specific clinician training) | Level of cultural security in health care setting |
| None | None |
| **Engagement of the whole practice** (closest: Institutional culture/motivation to change) | System allows followups of same patient, Adequate health staff levels |
| Specific clinician training | **Community visits difficult to co-ordinate** |
| None | Specific clinician training |
| None | None |
| General/specific clinician training | None |
| None | None |
| None | None |
| None | None |
| None | Staff turnover, Knowledge of CWC, Knowledge of prevalence, Knowledge of cultural security, Ability to confirm presence of CWC |
| Specific clinician training | None |
| None | Staffing levels, **compliance to required treatment** |
| None | Weight of competing medical demands |
| None | Adequate health staffing levels, weight of competing medical demands |
| None | Perceived as priority |
| **Posters of guidelines** (closest: easy to find guidelines), **reminders** (closest: easy to find guidelines), easy to understand guidelines | Perceived as priority, adequate health staff levels, **patient case complexity**, **insufficient resources** (closest: weight of competing medical demands, adequate health staff levels) |
| None | None |
| **Follow ups by phone**, level of cultural security in health care setting, system allows followups of same patient | Adequate health staffing levels |
| None | None |
| None | None |
| Adequate health staffing levels, weight of competing medical demands, General/specific clinician training, **mentoring** (closest: specific clinician training), **recognition for excellence (e.g. in treatment/knowledge)** (closest: Knowledge of CWC) | Knowledge of CWC/prevalence/cultural security, Adequate health staff levels, Weight of competing medical demands, Staff turnover |

Taken as a whole, these results suggest that the BN provides good coverage of the key determinants for any successful intervention (item 3 in the assessment list). It also provides evidence that the variables have been appropriately defined (the first part of item 4), as they frequently matched well with suggested facilitators and barriers. Items 3 and 4 were further validated via face validations of the model during development, which occurred at the beginning of every expert elicitation session. Table 6 shows a summary of model changes requested by the experts in each of these sessions. In the final two expert sessions, no new determinants were identified, only one determinant was renamed, and no changes were made to the possible states.

**Table 6:**
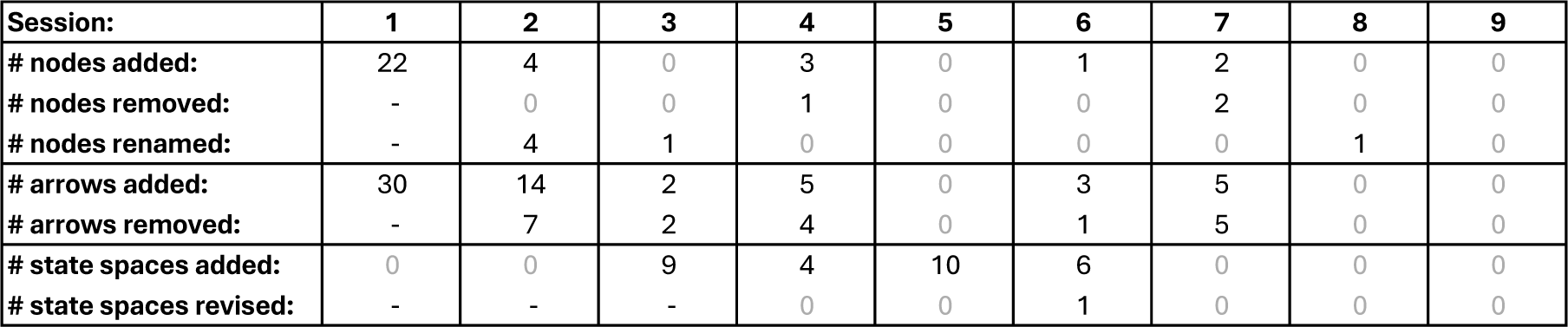
Changes in APPLE BN elements across sessions. The table summarises the changes in nodes, arrows and state spaces (i.e., the sets of possible states for each variable) of the APPLE BN throughout its development, from the initial expert session to subsequent elicitation and validation sessions. **Session 1:** No prior BN existed, so no elements (nodes, arrows or states) were removed. **Session 2:** Modifications included adding and refining nodes and arrows. **Session 3:** State spaces for nodes were introduced and considered for the first time. **Sessions 4-8:** Modifications continued for all BN elements, with the fewest changes in Session 8. **Final session:** No further changes were made, indicating the BN had stabilised.

The qualitative validations above support items 1 and 2. However, quantitative methods can also be applied, particularly for item 1. Good validation practice requires that evaluation data be separate from development data — for example, using k-fold cross-validation to separate training data from test data in each iteration. To ensure this separation, we performed cross-validation by validating a model developed from one subset of data (e.g., expert input) against a different subset (e.g., survey data) and vice versa. This provides a more robust and reliable model assessment. If the results are consistent across these validations, it strengthens our confidence in both the model and the underlying data sources.

To assess item 1, we need to check the causal influence on the outcome or other intermediate nodes for every determinant in the model, alone and in combination. Since the model here is primarily linear and independent, focusing on each determinant’s influence on the outcome in isolation is a reasonable approach. We begin by looking at the sensitivity of the outcome, **Appropriate management**, to all other nodes for both the expert BN and survey BN as shown in Table 7.^3^ There is shared information between the two BNs in this case, which is the assumed causal structure; hence, this is a cross-validation of the *parameters*. The sensitivity is based on both MI (measuring the *association* between variables) and CI (measuring the *causal* or *interventional* power, assuming the model structure is causally correct and sufficient). We can see substantial agreement in the rankings between the two BNs, with a rank correlation between the two node orderings over CI of 0.92 (and over MI of 0.91).

**Table 7:** Sensitivity of **Appropriate management** to other variables. These tables show the relative importance of variables and their roles in shaping the outcome by summarising the sensitivity of **Appropriate management** to each. The left table shows variable sensitivities from the expert BN, the right from the survey BN. Sensitivity is measured using MI and CI, and the columns marked ‘MI %’ and ‘CI %’ show these as a percentage of their maximums. (See Section 2.2 for measure descriptions.) Notably, even small values for MI or CI can still represent meaningful influences. For instance, in the expert BN, the variable **Easy to find Guidelines** has a CI of 0.2%, but this corresponds to a 5% change in the probability of good **Appropriate management** (from 25% to 30%).

| Variable | MI | MI % | CI | CI % |
| --- | --- | --- | --- | --- |
| Ability to confirm presence of CWC | 0.32385 | 21.6 | 0.27310 | 17.2 |
| Knowledge of cultural security | 0.08371 | 5.6 | 0.07138 | 4.5 |
| Ask about presence of CWC | 0.06489 | 4.3 | 0.04281 | 2.7 |
| Level of cultural security in health care setting | 0.03380 | 2.3 | 0.03403 | 2.1 |
| Knowledge of guidelines | 0.02080 | 1.4 | 0.01738 | 1.1 |
| Perceived as priority | 0.01214 | 0.8 | 0.00889 | 0.6 |
| Motivation to provide culturally secure care | 0.05753 | 3.8 | 0.00871 | 0.5 |
| Knowledge of CWC | 0.00650 | 0.4 | 0.00563 | 0.4 |
| Easy to find Guidelines | 0.00276 | 0.2 | 0.00390 | 0.2 |
| Easy to understand Guidelines | 0.00349 | 0.2 | 0.00374 | 0.2 |
| System allows followups of same patient | 0.00381 | 0.3 | 0.00372 | 0.2 |
| Confidence in treating CWC | 0.01306 | 0.9 | 0.00292 | 0.2 |
| Staff turnover | 0.00160 | 0.1 | 0.00253 | 0.2 |
| Specific Clinician Training | 0.00095 | 0.1 | 0.00141 | 0.1 |
| General Clinician Training | 0.00083 | 0.1 | 0.00104 | 0.1 |
| Peer perceptions and behaviour | 0.00017 | 0 | 0.00014 | 0 |
| Weight of competing medical demands | 0.00011 | 0 | 0.00012 | 0 |
| Peer perceptions and behaviour in inner setting | 0.00005 | 0 | 0.00006 | 0 |
| Adeq. Health staff levels | 0.00002 | 0 | 0.00003 | 0 |
| Knowledge of prevalence | 0.00026 | 0 | 0.00002 | 0 |
| Institutional culture/motivation to change | 0.00001 | 0 | 0.00001 | 0 |
| Peer perceptions and behaviour in outer setting | 0.00000 | 0 | 0.00001 | 0 |
| Pressure from families | 0.00000 | 0 | 0.00000 | 0 |
| Pressure from public/media/social media | 0.00000 | 0 | 0.00000 | 0 |

Table 7: Sensitivity of **Appropriate management** to other variables.
| Variable | MI | MI % | CI | CI % |
| --- | --- | --- | --- | --- |
| Ability to confirm presence of CWC | 0.26569 | 17.3 | 0.17429 | 11 |
| Knowledge of cultural security | 0.10310 | 6.7 | 0.14660 | 9.2 |
| Ask about presence of CWC | 0.10428 | 6.8 | 0.03662 | 2.3 |
| Level of cultural security in health care setting | 0.03157 | 2.1 | 0.03551 | 2.2 |
| Knowledge of guidelines | 0.12243 | 8 | 0.09102 | 5.7 |
| Perceived as priority | 0.00536 | 0.3 | 0.00510 | 0.3 |
| Motivation to provide culturally secure care | 0.08374 | 5.5 | 0.04571 | 2.9 |
| Knowledge of CWC | 0.00186 | 0.1 | 0.00079 | 0.1 |
| Easy to find Guidelines | 0.02597 | 1.7 | 0.02784 | 1.8 |
| Easy to understand Guidelines | 0.00884 | 0.6 | 0.00993 | 0.6 |
| System allows followups of same patient | 0.00032 | 0 | 0.00192 | 0.1 |
| Confidence in treating CWC | 0.02992 | 1.9 | 0.00331 | 0.2 |
| Staff turnover | 0.00538 | 0.4 | 0.01398 | 0.9 |
| Specific Clinician Training | 0.00003 | 0 | 0.00007 | 0 |
| General Clinician Training | 0.00004 | 0 | 0.00012 | 0 |
| Peer perceptions and behaviour | 0.00051 | 0 | 0.00062 | 0 |
| Weight of competing medical demands | 0.00004 | 0 | 0.00004 | 0 |
| Peer perceptions and behaviour in inner setting | 0.00005 | 0 | 0.00005 | 0 |
| Adeq. Health staff levels | 0.00000 | 0 | 0.00000 | 0 |
| Knowledge of prevalence | 0.00004 | 0 | 0.00009 | 0 |
| Institutional culture/motivation to change | 0.00001 | 0 | 0.00002 | 0 |
| Peer perceptions and behaviour in outer setting | 0.00005 | 0 | 0.00006 | 0 |
| Pressure from families | 0.00000 | 0 | 0.00000 | 0 |
| Pressure from public/media/social media | 0.00000 | 0 | 0.00001 | 0 |

Overall, the sensitivities in the tables, particularly towards the bottom (generally involving variables towards the top of the BN structurally), may be lower than what we may expect. This is likely due to the notable uncertainty in the local relationships for both BNs. This appears to be particularly the case with the expert BN, with 5 variables having greater than 1% CI with the outcome, while in the survey BN, 7 of the variables do. The variables towards the bottom of both tables have sensitivities near 0. Since these may artificially inflate the rank correlations between the BNs, we can look at the rank correlation between the 10 variables that rank highest in the expert BN. Doing so yields a lower but still strong 0.75 rank correlation over CI (and 0.76 over MI). While the rank correlations over CI and MI are nearly identical, thetwo measures have some apparent differences. For example, **Motivation to provide culturally secure care** has a notably weaker CI than MI in both BNs, while other variables such as **Ability to confirm presence of CWC** and **Knowledge of cultural security** have mixed measurements.

Table 8 shows the MI of every variable with the **Appropriate management** outcome in the raw survey data, and compares these with the MIs in both the expert BN and survey BN. The table is ordered by the MI in the raw survey data. We can see that the absolute value of the MI is quite different in the survey data compared to the two BNs. This is expected, particularly for nodes increasingly far from **Appropriate management** in the BN causal pathways. Again, the relative rankings of the MIs are likely to be more informative as to how well the survey data supports the validity of the expert BN and, to a lesser extent (due to shared information), the survey BN. The rank correlation between the survey data MI and expert BN MI ranks is 0.50, suggesting moderate support for the expert BN. The rank correlation with the survey BN is somewhat lower, at 0.45.

**Table 8:**
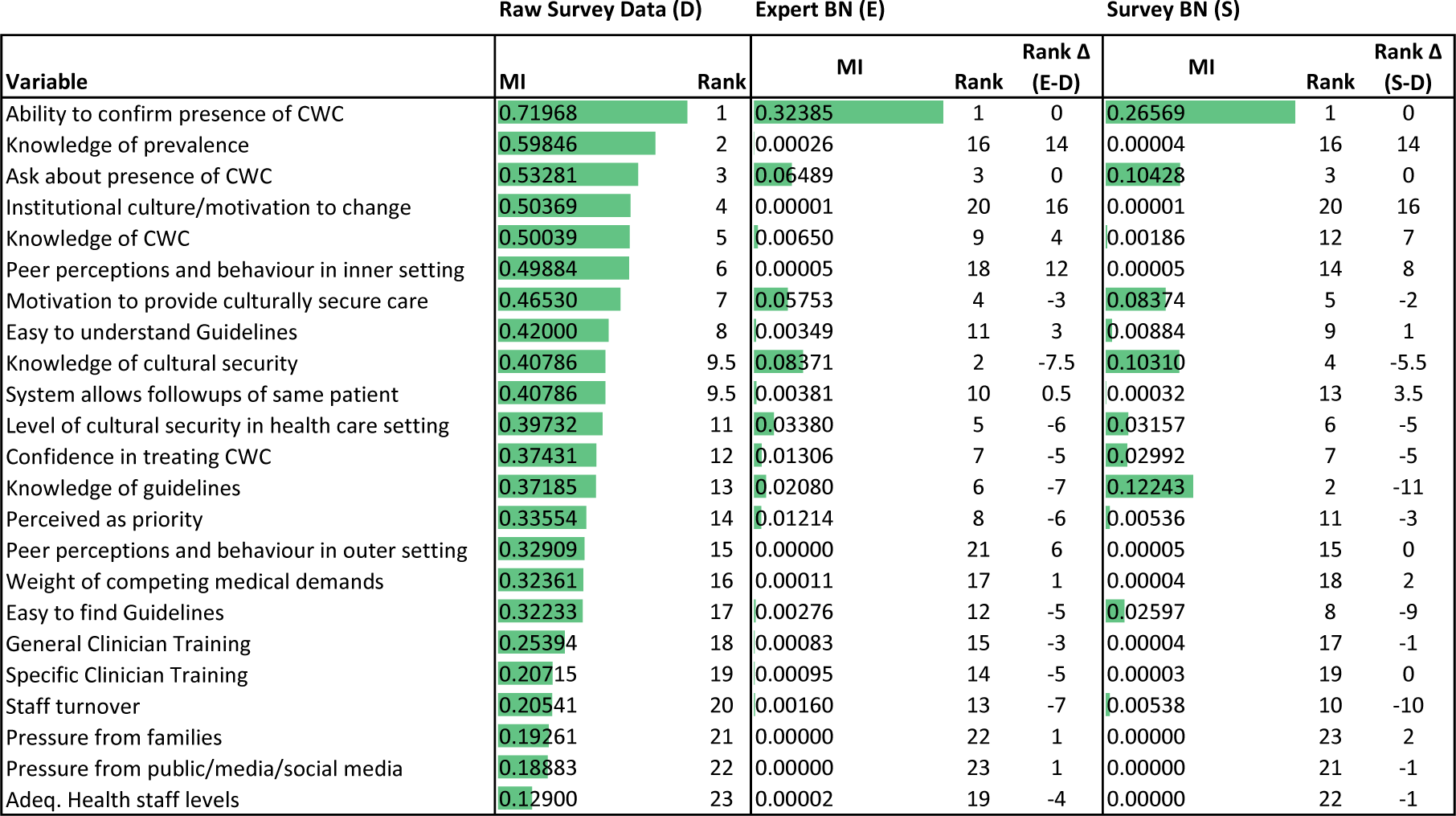
Each variable’s MI (and MI rank) with the outcome. This table presents the Mutual Information (MI) and corresponding rank of each variable in relation to Appropriate Management across three contexts: (D) the raw survey data, (E) the expert BN, and (S) the survey BN. Since the raw survey data lacks a causal structure, CI is not included in this comparison. For both the expert and survey BNs, the table also provides the difference between the MI rank in the BN and the rank derived from the raw survey data. This highlights any discrepancies in the importance of variables as interpreted through the BN models versus the raw data.

The table also shows the rank differences between the two BNs and the survey data. Focusing on the expert BN^4^, the average absolute rank difference is 5 (compared to an expectation of 7.7 for randomly ordered ranks). Three variables are particularly large outliers relative to that average: **Institutional culture/motivation to change** (rank difference of 17), **Knowledge of prevalence** (rank difference of 14), and **Peer perceptions and behaviour in inner setting** (rank difference of 11.5). Omitting those 3 outliers gives a strong rank correlation of 0.83; the average absolute rank difference also reduces to 2.7 against an expectation of 6.7 for randomly ordered ranks over the remaining variables. This suggests good agreement over the remaining variables. Of course, agreement does not necessarily mean they both represent the true situation (the collected data and the model could both be wrong). However, disagreement necessarily implies that either the survey data or the expert BN does not represent reality.

The considerable disagreement over these 3 variables clearly implies that the survey data or the expert BN do not represent the actual influence of these variables. If we assume the survey data is the better fit, we can examine the BN to see what adjustments might need to be made. **Institutional culture/motivation to change** and **Peer perceptions and behaviour in inner setting** are strongly related conceptually (and, indeed, were made parent and child in the BN). Their influence in the BN is solely via **Perceived as a priority**, which is only moderately related to the outcome in the data and the expert BN. Hence, further causal pathways may need to be added to represent the impact of these two nodes on the outcome. Alternatively (and since we can only examine MI, i.e., association, from the data) there may be further *associations* in the BN that are needed, such as back paths from these 2 nodes to the outcome.^5^ **Knowledge of prevalence** also only affects the outcome via **Perceived as a priority**. It does have additional associations through back paths, all via **General clinician training**, though those back paths are very weak. This similarly suggests it may act via other causal pathways, potentially **Knowledge of CWC**, given the importance of this node in the expert BN, or via a more direct relationship with **Ask about presence of CWC** (even more strongly ranked by the survey data and the two BNs).

### 3.3 Generalised workflow

An expert in Bayesian networks oversaw the development of the APPLE BN alongside clinical experts with a strong knowledge of implementation science concepts. Our team also had ample time and existing knowledge to tailor the design of the BN, the survey and the analysis to our implementation. It would be impractical to assume that every implementation program would have access to similar resources (particularly BN expertise). Hence, we developed a generalised workflow that teams with no specialised background knowledge in BNs, implementation science or statistical analysis can adopt.

The generalised workflow builds directly upon the development processes used for APPLE BN. It provides a structured method for integrating a causal analysis into the design of an implementation program that anyone can follow. One key extension of the process used for APPLE BN is that it leverages the benefits provided by determinant frameworks early, incorporating them from the beginning of the workflow. An outline of the workflow is presented in Figure 2. Several roles and teams may be involved in the workflow, but here, we focus on the analysis team and their role within the broader implementation team. The workflow begins with the implementation team identifying the desired outcomes that the implementation aims to improve. Once these are chosen, the team must specify metrics for measuring the outcomes. The metrics allow the success of an implementation to be evaluated. While outcome metrics are not strictly needed to develop the causal BN or for its use in planning the implementation, identifying them early allows it to be developed with the outcomes and their measures in mind.

After selecting the desired outcomes, the implementation team selects a determinant framework. A suitable determinant framework will include most or all the determinants that notably influence the outcome.^6^ The CFIR is a good general candidate framework for health implementations, as the broad and fine-grained domains of the CFIR are comprehensive. With a determinant framework selected, the analysis team will then identify likely determinants. The framework assists in this process by mapping out the domains that should be considered without specifying the exact determinants. Applicable methods include elicitation (e.g., nominal group technique) or thematic analysis of interviews.

Once a candidate list of determinants has been identified, the analysis team uses it to build the causal BN. This process can be guided by literature or conducted as an elicitation process (Mascaro et al., 2024b describes such a process). The purpose is to identify how the determinants interact with each other to influence the implementation outcomes. At a minimum, variables within the causal BN will consist of the desired outcomes and the identified determinants. However, as the causal mechanisms linking these variables are elaborated, new variables will likely be needed to provide a useful account of the causal processes governing an implementation. These new variables may be convenient intermediate variables (such as a summation variable) but may also be new determinants. After the causal BN structure has been developed (including variables and causal arrows), the analysis team must specify each variable’s possible states. This, too, may lead to further changes in the BN or its determinants.

The next step is the parameterisation of the causal BN — specifically, *a* qualitative parameterisation, which captures the direction and quality of changes in probabilities, not the precise quantitative changes (as described in Mascaro et al., 2024a). This parameterisation provides the causal BN with behaviour — specifying whether a determinant is an enabler, barrier or has some more complex relationship with other determinants and the outcome. This allows determinants to be assessed and compared in terms of their influence over the outcomes. It is also possible to assess determinants based on the structure of the causal BN alone, but this results in significantly less accurate estimates of influence.

With a qualitatively parameterised causal BN, the analysis team can evaluate the model with experts. If the evaluation provides enough confidence in the model’s behaviour, and if the implementation is low risk, the analysis team may choose to end the analysis at this point, rank the determinants by their likely impact on the outcomes, and use this ranking to inform the design of the implementation program directly. Alternatively, the analysis team may decide that a more representative parameterisation is warranted that would involve collecting data through observations or structured surveys. The qualitatively parameterised causal BN can guide the design of this data collection. If data is collected, additional validation can be done on both the qualitatively and data-parameterised causal BNs.

Finally, the information needed at each stage of the development of the parameterised causal BN can be obtained via formal elicitation processes, for example, one in which a facilitator elicits information from experts (i.e., those that hold that information) using a formal protocol (such as the Delphi technique; Linstone and Turoff, 1975), or otherwise obtained through an informal or unstructured information gathering process. The workflow presented here allows for both.

## 4 Discussion

A causal Bayesian network model was developed to map and quantify how contextual and other factors might affect the detection and management of chronic wet cough in Indigenous children in primary health care settings. APPLE BN provides a method for understanding and quantifying 1) the mechanisms through which implementation strategies operate and 2) the effect of individual factors on each other and implementation outcomes. Hence, the BN can be used to plan implementation in Phase 2 of the Implementation Roadmap (Barwick, 2023), or study implementation mechanisms. A survey linked to the BN could be applied at individual clinics to determine the local status of contextual factors, thereby allowing an estimation of the proficiency of local clinicians to detect and correctly manage chronic wet cough. The BN can predict the effect of change (of a specified magnitude) in one variable on another and thereby allows the implementation team to select strategies most likely to bring about such change. To our knowledge, this is the first description of a methodology to statistically quantify the relationships between various contextual, and other factors that might influence the uptake of evidence into clinical practice and allows the effects of different implementation strategies to be explored *in silico*.

The BN provides a visual and statistically robust method for studying implementation mechanisms, highlighted as an important area in implementation research (Vejnoska et al., 2022). Lewis et al. defined a mechanism as “a process or event through which an implementation strategy operates to affect one or more implementation outcomes” (Lewis et al., 2020). They advocated standardising terms in this area by clearly defining preconditions, strategies, determinants, mediators and proximal and distal outcomes. Our BN quantifies how determinants, mediators and moderators are interrelated and how implementation strategies might influence outcomes through complex interactions with these factors. The BN allows for the visual identification of mediators (a variable that transmits the effect of an independent variable on a dependent variable), moderators (variables that change the magnitude of the relationship between other variables), and mechanisms by describing the exact series of steps through which change occurs as outlined by Kazdin (2007). The BN does not contain any preconditions (factors needed to activate implementation mechanisms (Lewis et al., 2020)). We were aware that clinical practice guidelines supporting the evidence around the management of CWC did exist. Clearly, recognising such guidelines by a health care institution would be a precondition to such guidelines being followed. A particular strength of the BN is that it allows the identification and testing (through Bayesian modelling) of multilevel mediators (Williams, 2016).

The BN is not the first attempt to visually map out how an implementation project’s determinants, mechanisms, strategies and outcomes interrelate. The Implementation Research Logic Model also graphically depicts the connections between these factors (Smith et al., 2020) and, unlike our BN, includes a description of strategies and outcomes. Whilst the Implementation Research Logic Model is beneficial for planning implementation projects, it is not a causal model that allows the mathematical modelling of causal relationships within a system. Specifically, it does not describe how determinants in a system relate to each other and does not allow the strength or effects of relationships to be quantified. Hence, BNs and logic models have different but complementary roles in implementation science.

The BN is also not the first attempt to develop systematic methods for selecting the core components of an implementation process. The process of implementation mapping was developed as a systematic process for planning and selecting implementation strategies (Fernandez et al., 2019). However, unlike implementation mapping, a BN provides a quantifiable method for predicting the likely effect of changes to a single or multiple factors targeted by implementation strategies, thereby adding scientific rigour to the strategy selection process. BNs can be used in conjunction with implementation mapping when planning implementation.

Whilst the BN was designed to include relevant CFIR determinants, it did not include the characteristics of individuals involved in the implementation process. Hence, the BN cannot predict how well a strategy will be implemented. One would expect larger effects when the implementation team is skilled and effective. However, the BN does factor in important distal implementation outcomes (Proctor et al., 2011) like acceptability, adoption, and penetration, and provides local context through the survey that feeds into the BN. For this specific BN, which focuses on the uptake of evidence-based guidelines on the management of chronic wet cough, adoption of the guidelines have already been shown to result in improved clinical outcomes (Laird et al., 2021).

A potential weakness of our BN is that it was initially informed by the original CFIR, which was revised whilst the BN was being developed. However, the original CFIR is a comprehensive, widely used and recognised determinant framework in implementation science, and its core structure has not changed with the recent revision. It is worth noting that the model is amenable to updates and changes, as the core structure will often remain unaltered as new determinants are added — this pattern being already set during its evolutionary development process.

The BN has been validated by comparing expert estimates with independently collected survey data. This has provided promising evidence of the BN’s ability to give well-calibrated and useful probabilistic estimates of the effects of determinants in isolation and in combination. Development of the BN also provided a secondary check of the design of the APPLE implementation, which proceeded in parallel, and this too indicated the BN’s validity and potential. However, the BN has yet to be used directly to inform or evaluate the design of an implementation program. Such a strong test of the BN’s performance and utility is currently being planned for a future stage of the APPLE program.

Finally, the BN focuses on the detecting and managing chronic wet cough in Indigenous children in primary care settings, a specific health condition in a selected health care environment. However, the Bayesian methodology incorporating a sound implementation science conceptual framework can likely be adapted to any other health condition and health care setting. We developed the generalised workflow (Figure 2) to provide a clear guide for applying the approach to any similar setting. The core causal analysis (yellow boxes) contains all elements and follows a nearly identical process to those used for APPLE BN. Later stages are identified (without elaboration) in the workflow by the grey boxes. We have left these later stages open, as they could use the causal BN in several ways: as examples, drawing on the information learned from the causal analysis, making direct use of the determinant rankings or using the causal BN itself as a model against which to test aspects of hypothetical programs prior to implementation or to guide the evaluation of implemented programs. However, it is worth outlining how we believe the design phase could be directly supported.

Figure 3 provides a potential approach to implementation program design based on the causal BN. The team uses the ranked determinants to identify intervention strategies, ensuring that strategies are relevant to (at least) the most important determinants. These strategies are then linked into the BN, via two alternative approaches: 1) an extension of the BN itself to incorporate these strategies, similar to how the determinants have been included in the BN; or 2) a set of simple ‘efficacy’ assessments that estimate the (independent) efficacy of each strategy on the root determinants (i.e., those without parents) in the BN. The latter approach would be particularly suitable for experts or implementers who may not be familiar with BN concepts. Once incorporated, the strategies can be assessed and ranked, much like the determinants themselves. Of course, many variations of an implementation process may use the strategies differently. These variations could all be assessed against the strategyaugmented causal BN. Depending on the nature of the variations and whether resource constraints apply, mathematical optimisation could be applied to identify a version of the process that maximises the probability of a successful outcome given the resources available (or, more generally, maximises the expected utility of the implementation).

For interventions intended to be implemented at multiple sites, it is critical to tailor the implementation process to each site’s circumstances and best strategy options. Hence, a second design stage might occur for individual sites, driven by a site assessment survey based on the developed causal BN. The survey would provide a rapid way to repeat the assessment (and potential program optimisation) across multiple sites, providing a way to scale implementation, while still providing site-sensitive implementations.

In conclusion, our BN provides an example of how Bayesian methods can be used to help select implementation strategies most likely to result in the uptake of evidence-based practices in health care settings, whilst also allowing the study of implementation mechanisms through the complex, multilevel interaction between determinants, mediators, moderators and mechanisms.

## Data Availability

Anonymised survey response data can be made available upon reasonable request to the authors.

## Footnotes

1 The use of ‘intervention’ has different connotations in implementation science and the philosophy of causation. Generally, we use the implementation science meaning of a larger strategy, program or action, but occasionally refer to interventions on individual variables where needed for clarity.

2 To isolate the influence of a selected parent, we assume independent uniform distributions for all parents and marginalise the parents to be removed. If we did not assume an independent uniform for every parent, their probabilities and dependencies would confound the analysis.

3 Sensitivity to **Appropriate management** itself is omitted, as it will be the maximum.

4 The following discussion applies almost equally well to the survey BN.

5 A back path is a path between two nodes that passes through a common cause, and is a source of confounding in statistical analysis when measuring the causal effect between the two nodes.

6 For example, if the desired outcome involves greater patient compliance with treatments, a framework that covers factors at home rather than within the clinic would be more appropriate.

7 In this appendix, ‘intervention’ refers to the manipulation of an individual variable.

8 CI tells us *how much more certain* we would become about the outcome if we intervened, not whether the outcome would be better or worse.

9 As an example, **Institutional culture** may have utilities associated with it that are just as significant as those for **Appropriate management**. However, we would not use this project’s resources to address that issue unless it contributed notably more to improving **Appropriate management** than other determinants. Ultimately, if **Institutional culture** was as significant on its own as **Appropriate management**, it should likely be the target of a separate project.

10 In some cases where resources are low or improvements are difficult, this may lead us to conclude that we have few to no feasible options.

11 ’+28%’ refers to the additive change in the probability, not the percentage improvement in the probability, which is 28%/42% or approximately 60%.

## Appendix A Parent strength of influence

**Figure 4:**
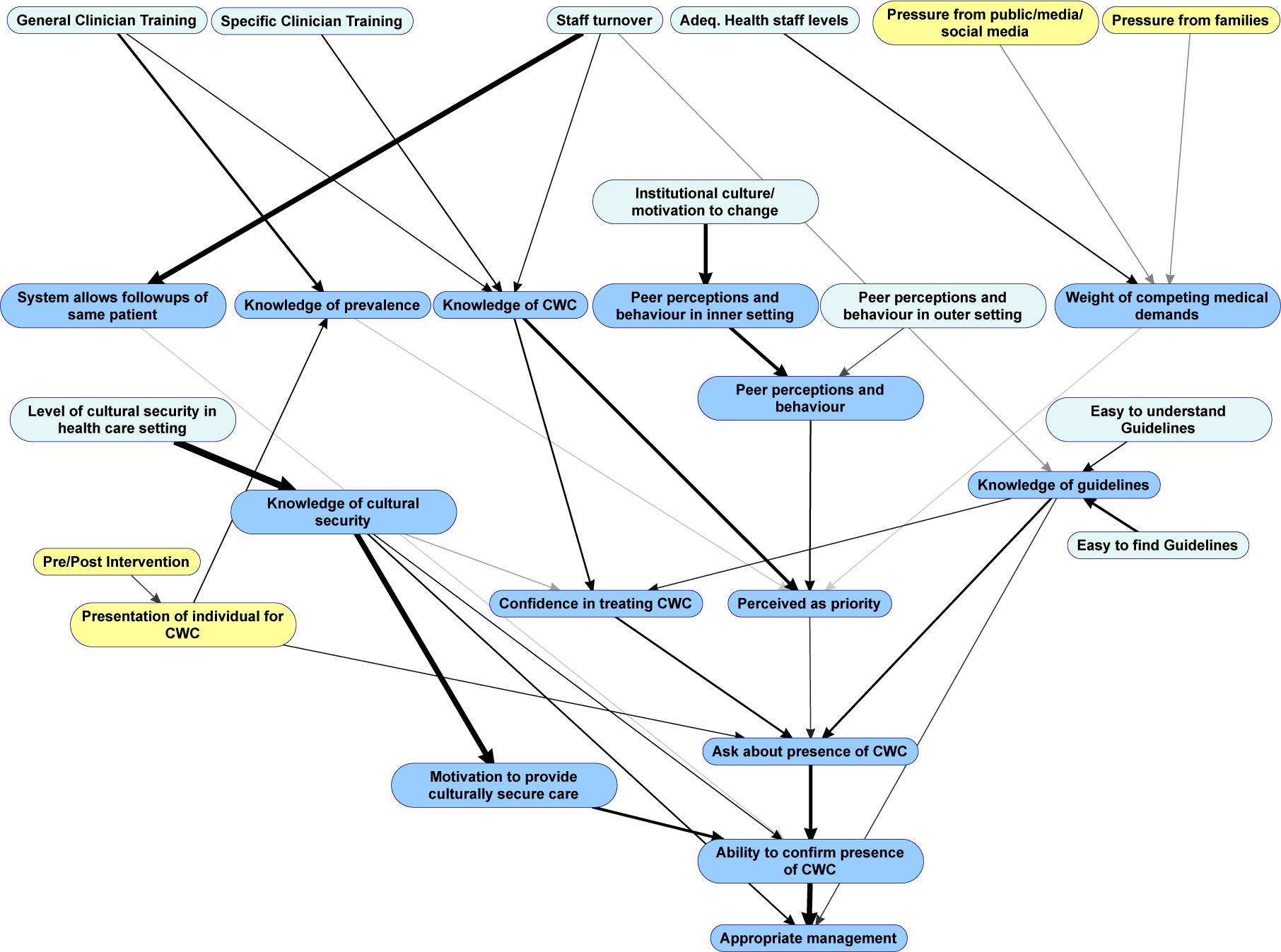
Strength of causal influences in the combined BN. The width and opacity of the arc indicates the strength of a parent node’s influence on a child node, measured as the mutual information between parent and child.

## Appendix B Example model usage

We provide 2 illustrations of how to use the APPLE BN to support implementation planning. The first illustration uses the model to create general recommendations that should work well for any site, and the second uses information about a site to create site-specific recommendations.

### B.1 Example 1: Creating general recommendations

We can generate an importance rankings of the BN determinants to obtain general recommendations for implementing an intervention.^7^ Assume for now that we will only look at determinants individually and not in combination. Some reasonable metrics for the importance ranking might be either mutual information (MI) or, better, (CI) (see Section 2.2 and Table 7). Determinants that rate high in causal MI are likely good candidates for strategies to target. However, these are measures of *information* and, hence, are still an indirect measure of what we would like to know for a determinant: i.e., How much do we expect the outcome to improve if we intervene on this determinant?^8^ This question has the structure of a decision, and can be modelled similarly to how we would model a decision in a Bayesian decision network (Howard and Matheson, 1981). The only difference here is we will do this for every node in the BN. After we attach values (or utilities) to different states of the outcome (and any other consequence), we test the best possible intervention on the determinant and then see how much the expected value improves. This static analysis is our current approach (compared to the interactive analysis we perform later).

We assign utility values only to the **Appropriate management** outcomes because that is our main focus. Other nodes might have their own utilities, but we do not consider them here.^9^ We also ignore the cost of interventions since costs can vary widely and are better addressed when deciding on specific site strategies. For **Appropriate management**, we assign the following values: 10 for **Good**, 0 for **Poor**, and 5 for **Moderate**, which is placed in the middle. Utilities are measured on an interval scale, so (aside from assignment order) only the **Moderate** assignment can affect our determinant ranking, and this effect is minor.

**Table 9:** Expected utilities from determinant interventions. (Middle column) The best expected utility (EU) achievable by intervening on a determinant in isolation. (Rightmost column) The difference between the best EU and the no-intervention EU.

| Node | Best EU | ΔEU |
| --- | --- | --- |
| Ability_to_confirm_presence_of | 7.04 | 2.19 |
| Knowledge_of_cultural_securit | 6.19 | 1.35 |
| Ask_about_presence_of_CWC | 6.05 | 1.2 |
| Level_of_cultural_security_in_ | 5.89 | 1.05 |
| Presentation_of_individual_for | 5.23 | 0.385 |
| Easy_to_find_Guidelines | 5.21 | 0.364 |
| Motivation_to_provide_cultural | 5.08 | 0.235 |
| Knowledge_of_guidelines | 5.07 | 0.225 |
| Perceived_as_priority | 5.07 | 0.222 |
| Confidence_in_treating_CWC | 5.05 | 0.206 |
| Easy_to_understand_Guidelines | 5.05 | 0.205 |
| Knowledge_of_CWC | 5.02 | 0.172 |
| Peer_perceptions_and_behaviour | 4.9 | 0.0512 |
| Specific_Clinician_Training | 4.88 | 0.0372 |
| Knowledge_of_prevalence | 4.88 | 0.032 |
| General_Clinician_Training | 4.87 | 0.0266 |
| Peer_perceptions_and_behaviour_inner | 4.87 | 0.0195 |
| Institutional_culture_motivati | 4.86 | 0.0137 |
| Peer_perceptions_and_behaviour_outer | 4.86 | 0.0131 |
| Pressure_from_public_media_so | 4.86 | 0.00901 |
| Weight_of_competing_medical_de | 4.86 | 0.00868 |
| System_allows_followups_of_sam | 4.85 | 0.00826 |
| Adeq_Health_staff_levels | 4.85 | 0.0062 |
| Pressure_from_families | 4.84 | 0.00164 |

We then compute the expected utility (EU) by setting each determinant, in turn, to its best state. The result is Table 9, which shows the expected utility associated with the intervention (Best EU), along with the improvement in the expected utility (Δ EU) against a baseline in which we do not intervene. This table can be used directly to design an implementation plan. For example, working down the table, we might note that **Ability to confirm presence** would have the greatest impact, but we have no direct way to intervene on it. By looking at the APPLE BN structure (Figure 1), we know many other determinants can affect it, so we leave it and move down the list. The following determinant is **Knowledge of cultural security** — of course, we cannot change that directly either, but we may have a training program that could be cost-effectively deployed. This may, therefore, become a central part of our implementation approach. We could continue working in this way down the list and consulting the APPLE BN structure until we no longer find improvements that would justify the resource cost.^10^

The value of this approach is that we target our interventions as close to our outcome as possible — but where we find no direct means to intervene, we can move further out to more distal determinants that can still have some or all the desired impact.

One limitation of this approach is that it requires one to manually work back and forth between the table and the BN structure. It also misses dependencies between determinants almost entirely. For example, if **Knowledge of guidelines** is only helpful if clinicians **Ask about presence**, then putting resources into improved guidelines without working to improve the inquiry rate may be wasteful. Similarly, if you can directly improve (say) the inquiry rate, it might mean that spending resources on other things (like general training) is no longer essential and is better directed at other areas. A light variant of the static approach above would be an interactive approach, i.e., using this table *interactively* while entering evidence into the BN. The basic process is the same: we generate the table from the BN with no evidence, identify the determinants we can target in order, but then enter our chosen interventions into the BN as we go, recomputing the table for the remaining determinants each time. Figure 5 shows an example of this over 4 steps for the APPLE BN, entering evidence for the best variable we can intervene on at each step and recomputing the expected utility of other determinants in the table each time. We see we can move the expected utility from 5.23 to 7.04 out of 10 (compare the Baseline EU above the table with the Best EU at the top of the table), which corresponds to an improvement in the probability of **Appropriate management** being **Good** of approximately +28%.^11^

Notably, a more intensive approach would be to use computational optimisation to select the determinants and the allocation of resources amongst them. An optimisation process would utilise the BN model, available data about the desired outcome, determinants that can be addressed to achieve the outcome, and constraints that must be satisfied within the clinic system, to suggest a set of strategies to achieve the desired outcome. Such optimisation is unlikely to produce significantly different general recommendations over the interactive approach, as much of the model’s non-linearity is already handled by the way the table is generated (in closest dependency order) and re-generated (i.e., dependent on the history of choices) each time. The interactive approach also allows one to interact and inspect the design of the implementation throughout the process, something formal optimisation techniques often do not make very convenient. Nonetheless, there are cases in which formal optimisation techniques would be the best approach, such as when there is a fixed budget that can be allocated to determinants in any proportion and we would like to identify the optimal allocation.

**Figure 5:**
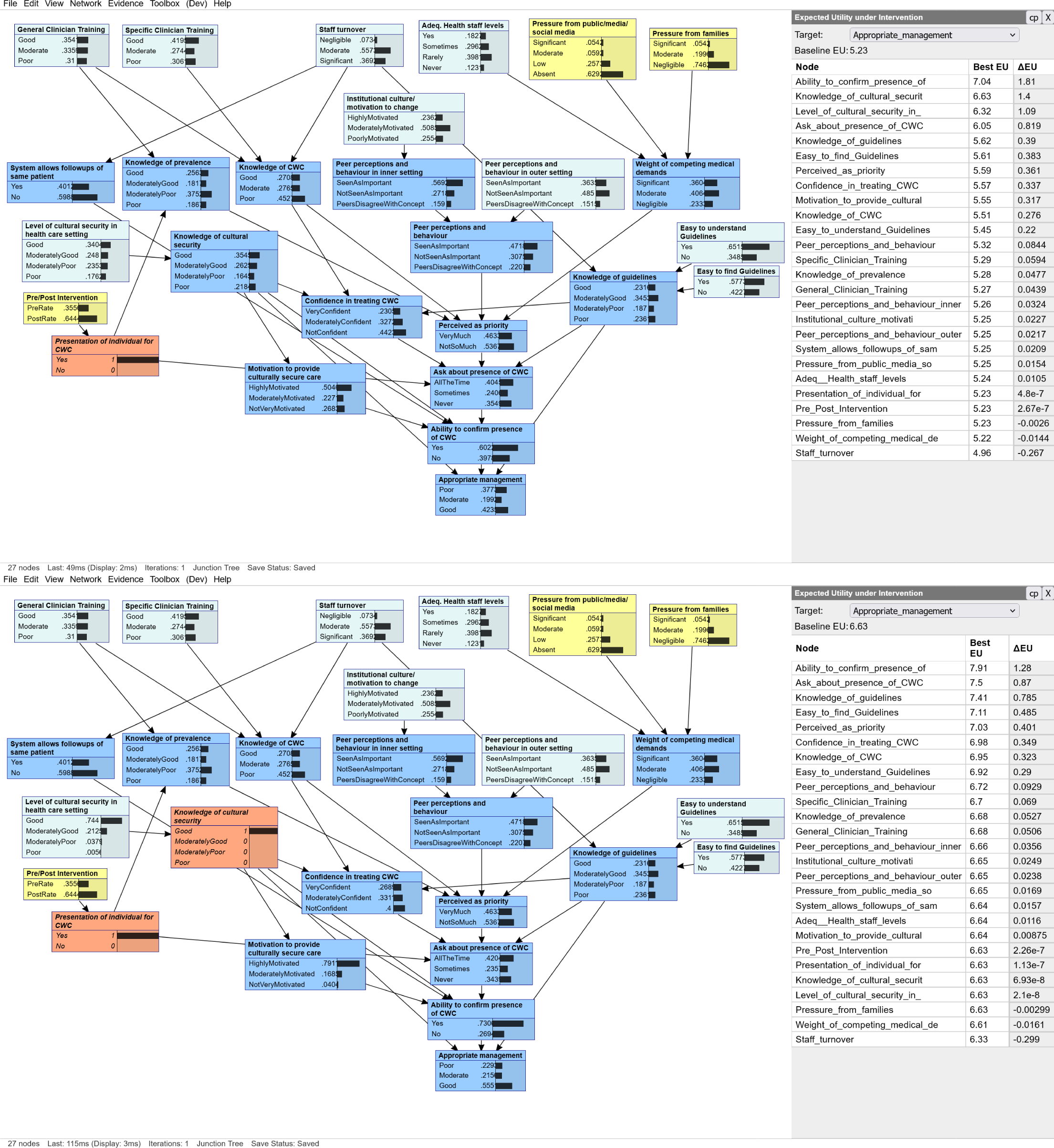
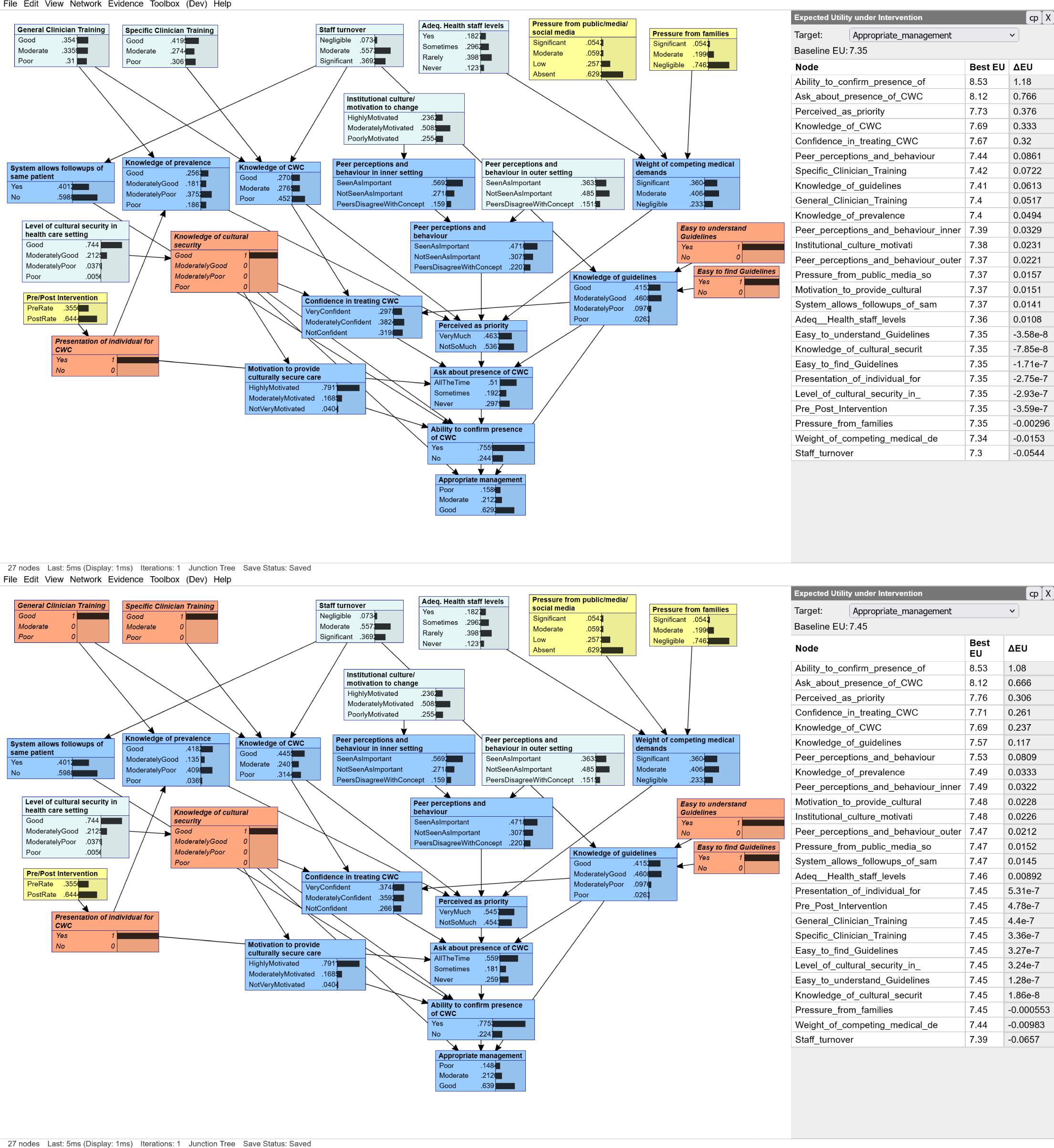
Illustration of the iterative method, in which the next best determinant is chosen dynamically based on determinants already chosen (and assumed to be already maximally improved). For example, Screen 1 shows no evidence (other than the assumption of Presentation of individual for CWC). The resulting ranking of nodes by impact on expected utility is visible in the table on the right of the screen. The next most influential node is chosen from this table and entered in Screen 2 (Knowledge of cultural security, the new orange node), which results in a new ranking over the nodes again. This is repeated in Screens 3 (with Easy to understand guidelines and Easy to find guidelines) and 4 (with General Clinician Training and Specific Clinician Training).

### B.2 Example 2: Creating site-specific recommendations

The above example deals only with general recommendations and does not consider that we typically have information about the implementing site. For example, if guidelines at a clinic are already accessible and easy to understand, we can direct resources to other determinants for that clinic.

The approach in this example builds on the static table method. First, we gather information about site determinants using the same APPLE BN survey that was used earlier for parameterisation and validation. However, unlike parameterisation, we can use any number of responses, even just one. We will call the distribution of responses for a determinant the “observed distribution” (even if it is based on a single response). Next, we calculate the best expected utility (EU) table as before, but this time compare it to a different baseline, namely the EU based on the observed distribution for the determinant. Finally, we rank all determinants by how much improving them would benefit the outcome compared to the observed distribution.

An example of doing this for a simulated case (modelled on one survey response that contained a mix of positive and negative answers) is given in Table 10. The collected observations are shown in the ‘Observation’ column. We can see that the areas the table suggests for improvement now begin with guidelines and perceptions of priority, with **Knowledge of cultural security** still high on the list but lower than before. The table suggests no improvements can be obtained from determinants such as **System allows followups** and **Motivation to provide culturally secure care**. Caution is required with this method when entering only a few responses. For example, the table also suggests no improvement can be obtained from **Ability to confirm presence of CWC** — perhaps this is true, but it is unlikely that there is no room for improvement here. When dealing with few responses, it would be better to assume that the responses are *approximate* and enter them as *uncertain* evidence rather than certain evidence. For simplicity, we omit this step from this example.

**Table 10:** As per Table 9, but with the current observation entered for each determinant in isolation.

| Node | Observation | EU given observation | Best ΔEU |
| --- | --- | --- | --- |
| Knowledge_of_guidelines | ModeratelyPoor | 5.46 | 0.964 |
| Easy_to_understand_Guidelines | No | 3.92 | 0.917 |
| Perceived_as_priority | NotSoMuch | 5.97 | 0.896 |
| Easy_to_find_Guidelines | No | 4.52 | 0.848 |
| Knowledge_of_cultural_security | ModeratelyGood | 5.31 | 0.504 |
| Knowledge_of_CWC | Moderate | 5.74 | 0.318 |
| Peer_perceptions_and_behaviour | NotSeenAsImportant | 5.95 | 0.247 |
| Confidence_in_treating_CWC | ModeratelyConfident | 6.32 | 0.218 |
| Knowledge_of_prevalence | ModeratelyGood | 5.93 | 0.102 |
| Peer_perceptions_and_behaviour_inner | NotSeenAsImportant | 5.28 | 0.0605 |
| Specific_Clinician_Training | Moderate | 5.01 | 0.039 |
| Peer_perceptions_and_behaviour_outer | NotSeenAsImportant | 4.76 | 0.0209 |
| Institutional_culture_motivati | ModeratelyMotivated | 4.77 | 0.0142 |
| Adeq_Health_staff_levels | Sometimes | 4.76 | 0.0101 |
| Weight_of_competing_medical_de | Moderate | 5.24 | 0.00666 |
| Pressure_from_families | Moderate | 4.78 | 0.00624 |
| General_Clinician_Training | Moderate | 5.01 | 0.00449 |
| Pressure_from_public_media_so | Moderate | 4.77 | 0.00253 |
| Level_of_cultural_security_in_ | Good | 4.76 | 0 |
| System_allows_followups_of_sam | Yes | 4.79 | 0 |
| Presentation_of_individual_for | Yes | 5.74 | 0 |
| Motivation_to_provide_cultural | HighlyMotivated | 5.98 | 0 |
| Ask_about_presence_of_CWC | AllTheTime | 7.44 | 0 |
| Ability_to_confirm_presence_of | Yes | 8.11 | 0 |

The above example gives a sense of how to generate recommendations at a specific implementing site. However, there are techniques for producing better recommendations and recommendations accompanied by explanations. For example, the interactive approach from Example 1 can also be applied here, and formal optimisation techniques can be applied too. However, additional techniques can take advantage of the causal structure of the model, working from the outcome backwards to causes, helping implementers understand how a specific site has come to have the outcome that it has now and, hence, where it is best to target resources. An explanatory approach that works backwards through causes is particularly useful since it can provide a robust understanding of how to plan an implementation, what to expect, and where to adjust if an implementation does not appear to have the desired effect. For example, the APPLE BN model might recommend improvements to perceptions of priority, explaining this will improve clinicians’ reliability in asking about and detecting chronic wet cough and suggesting that this can be achieved through improved training programs and improvements to institutional culture. This gives the implementer an understanding that allows them to adjust the implementation strategy, perhaps allocating resources to determinants that are closer to the outcome or to additional determinants that support the mediator. Ultimately, these are the robustly explained recommendations needed for complex implementation decisions.

## Appendix C APPLE BN Survey

### APPLE Study - Bayesian Practitioners Survey

Please complete the survey below.

Thank you!

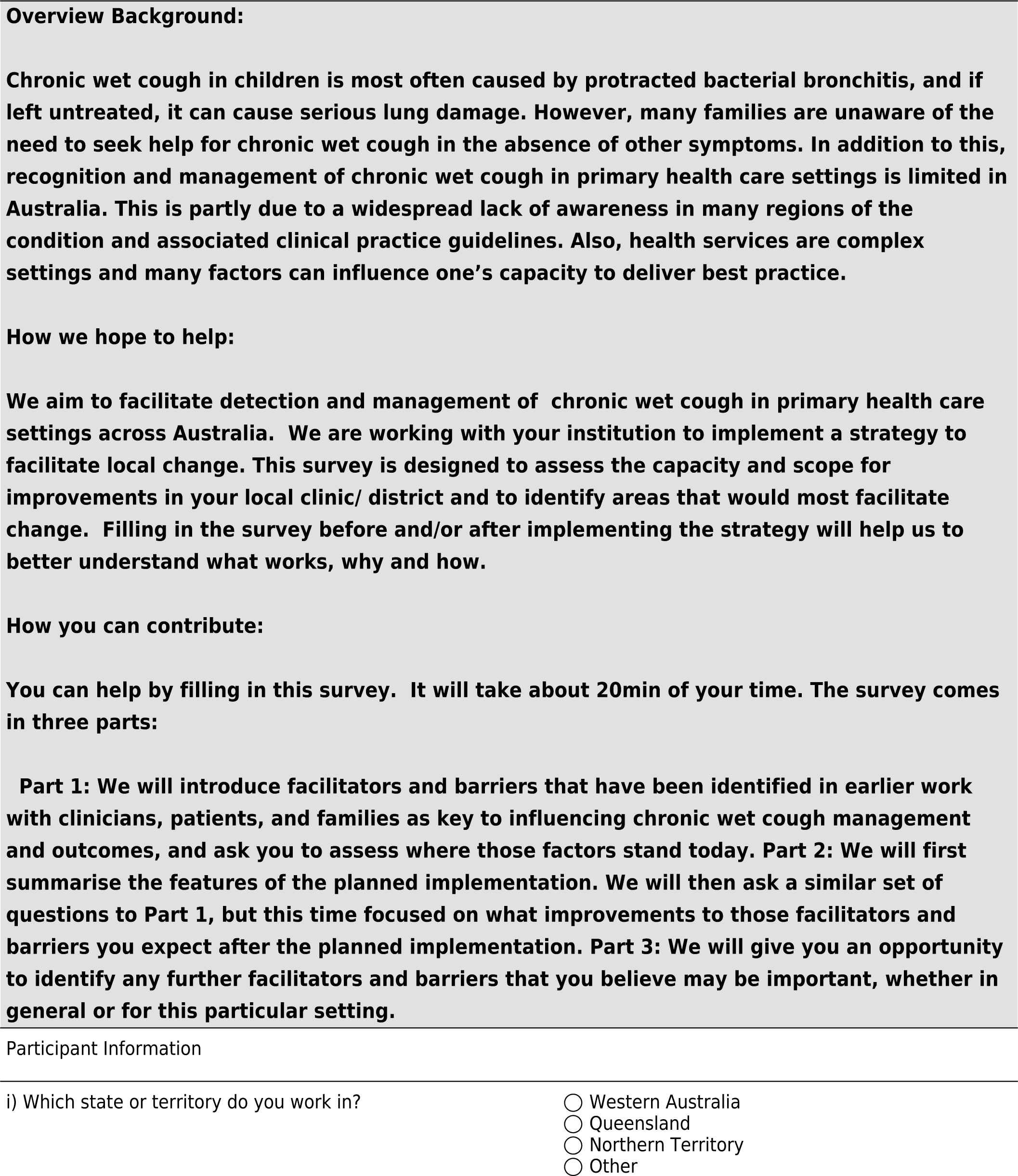
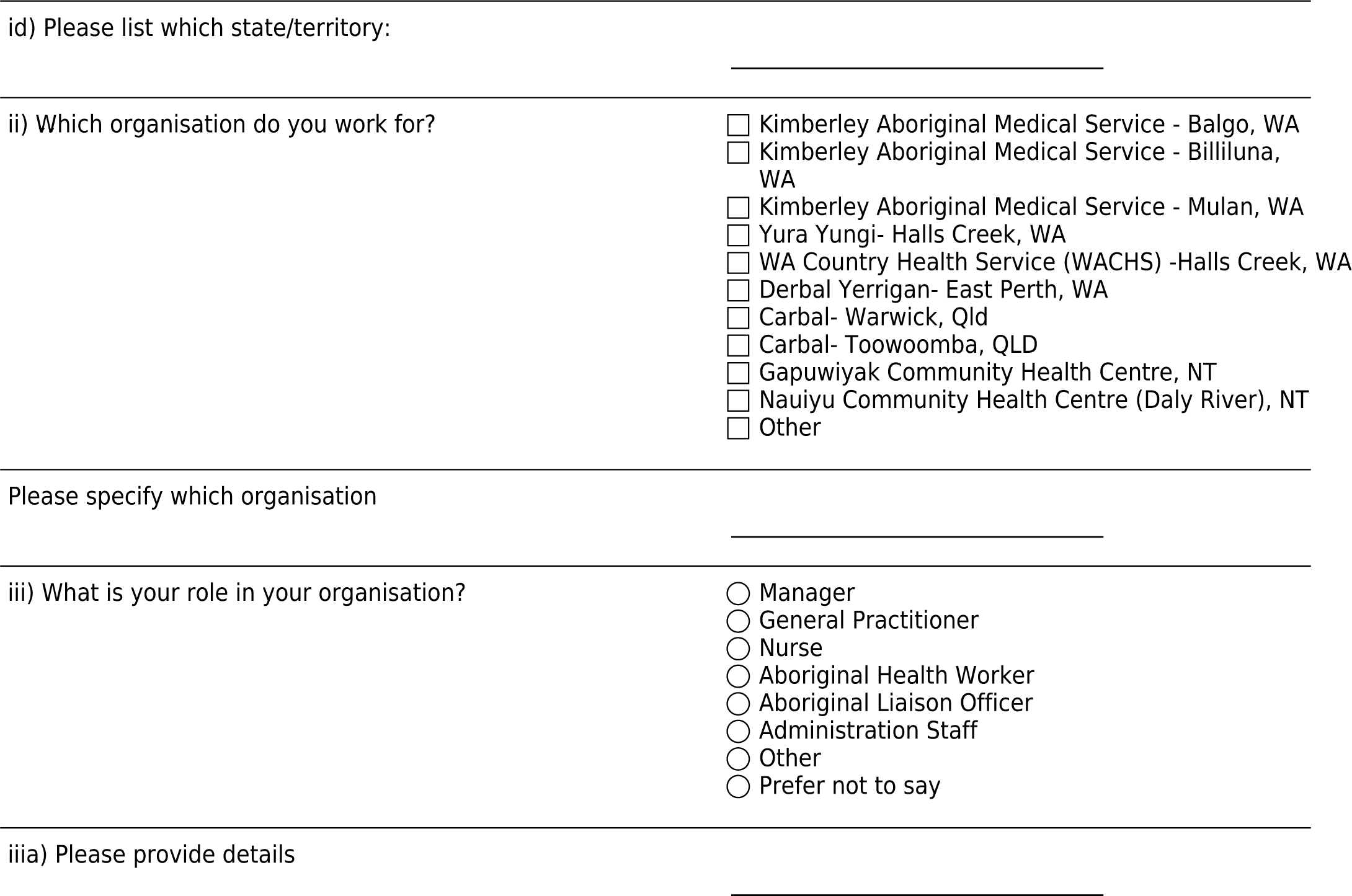

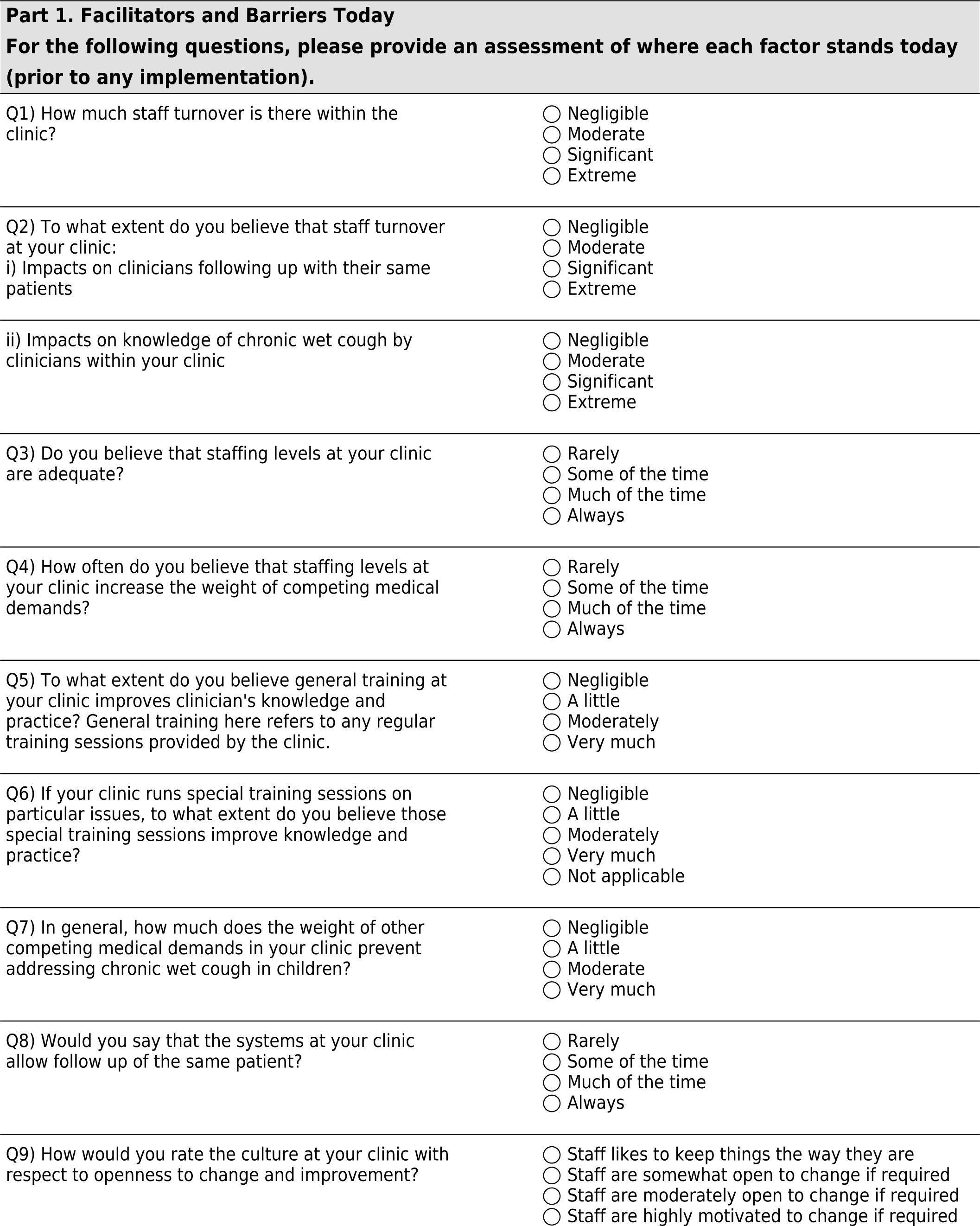
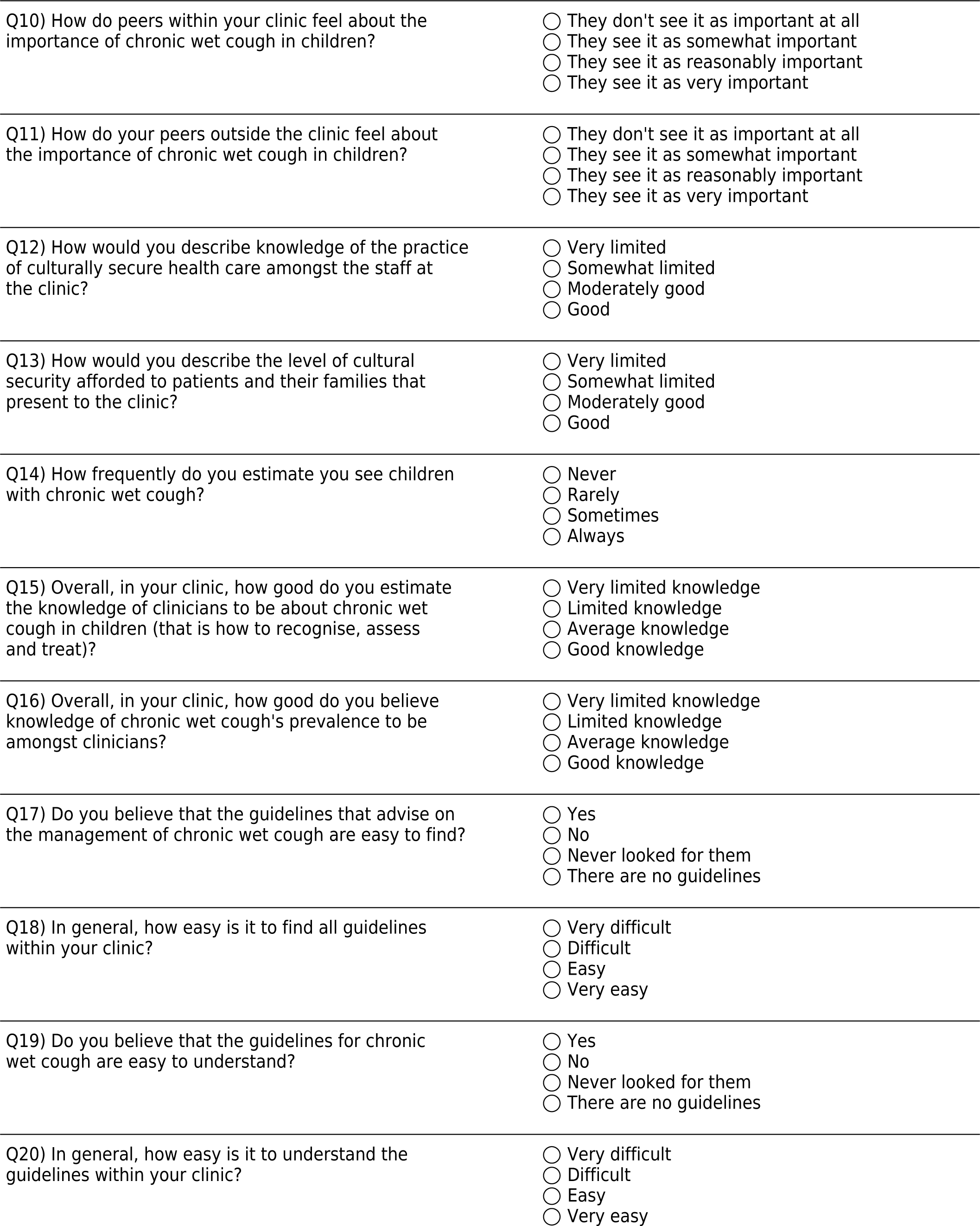
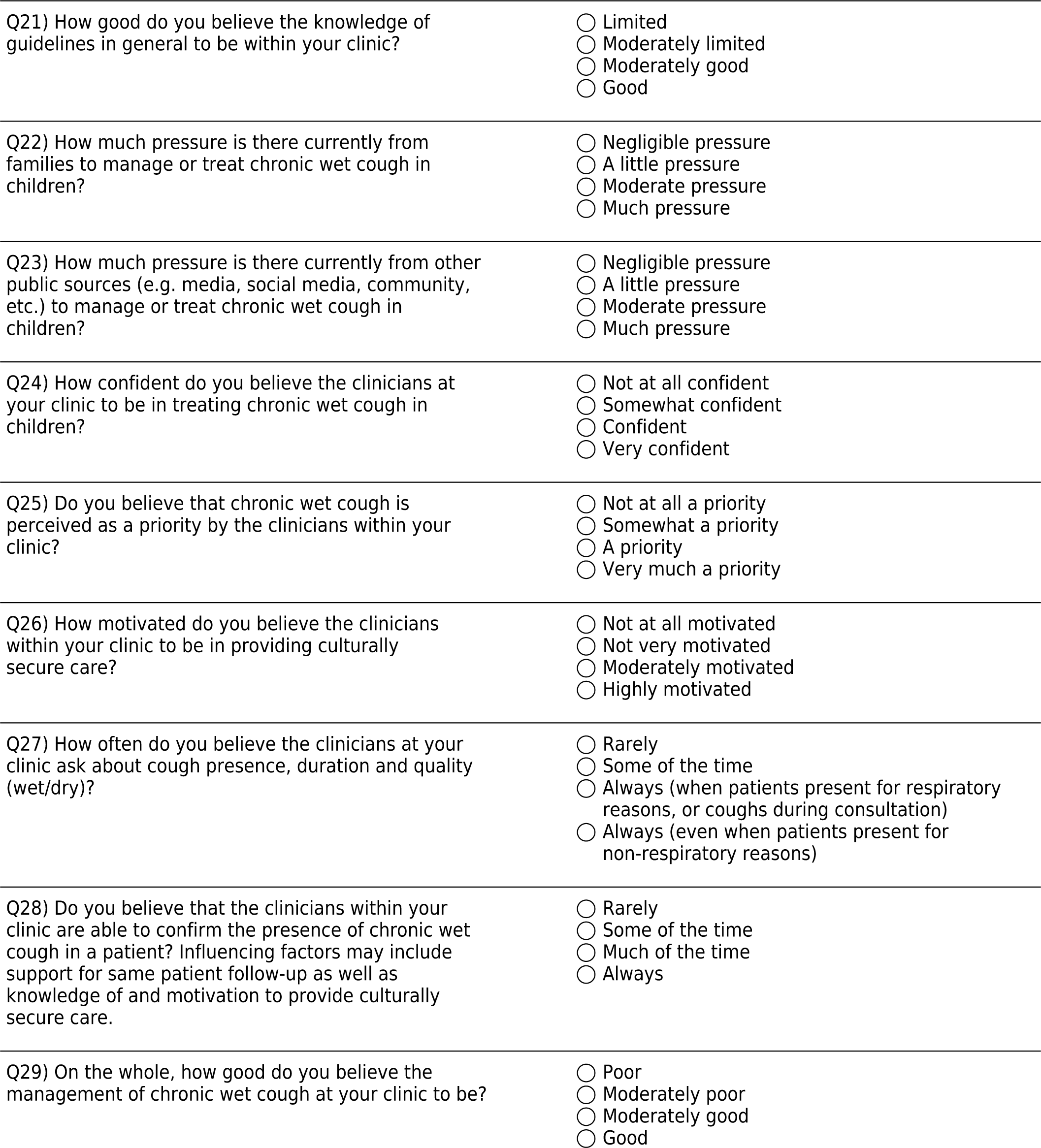

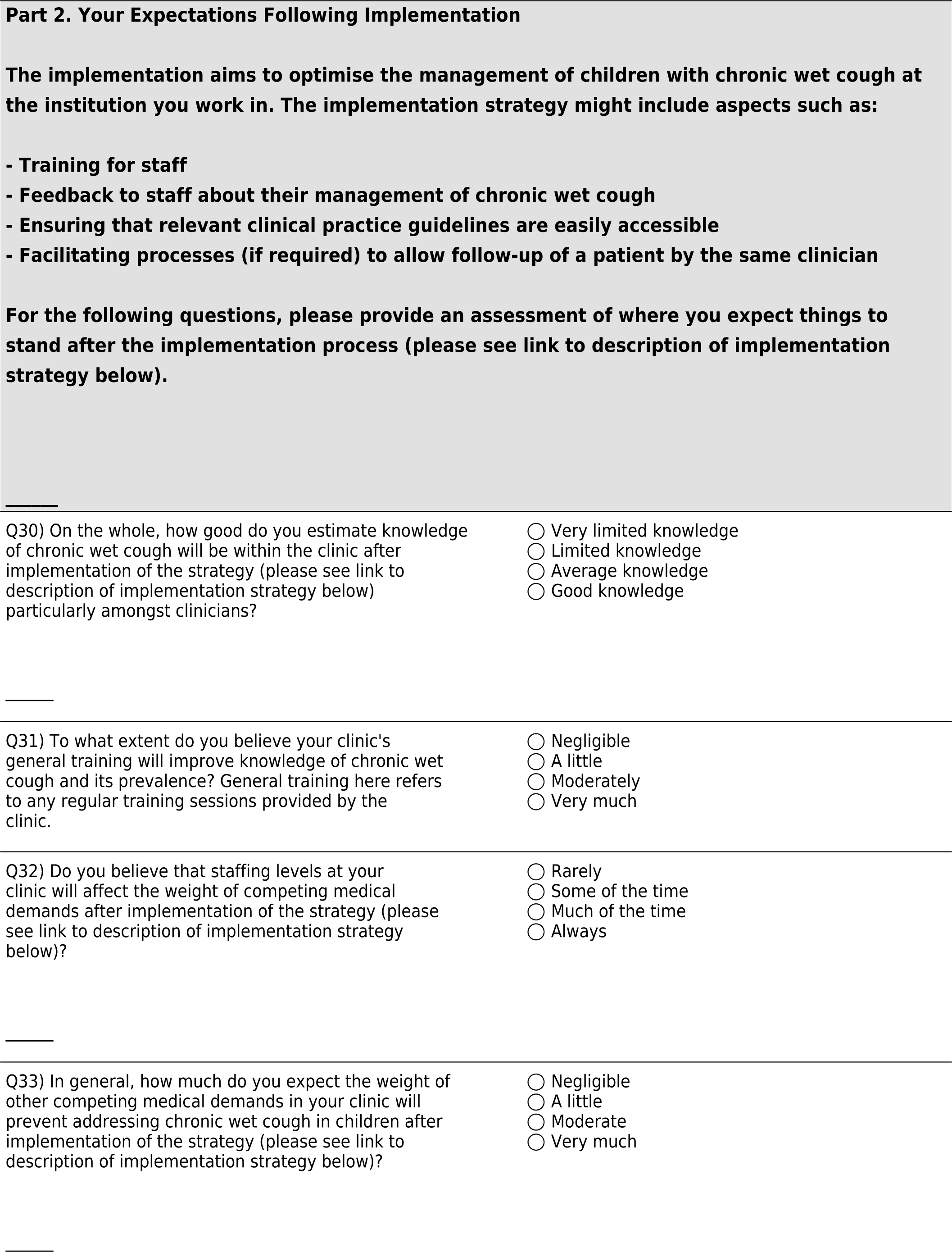
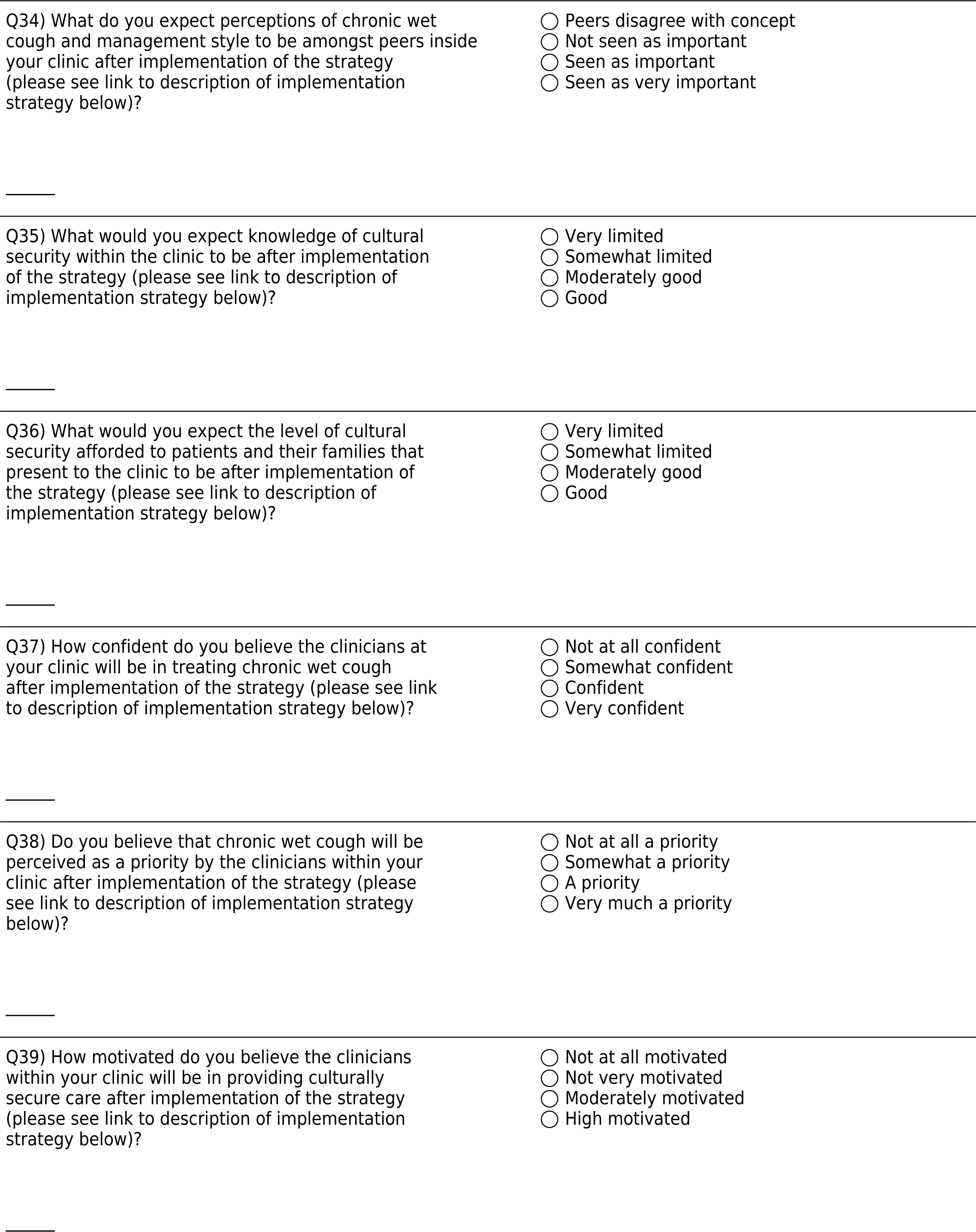
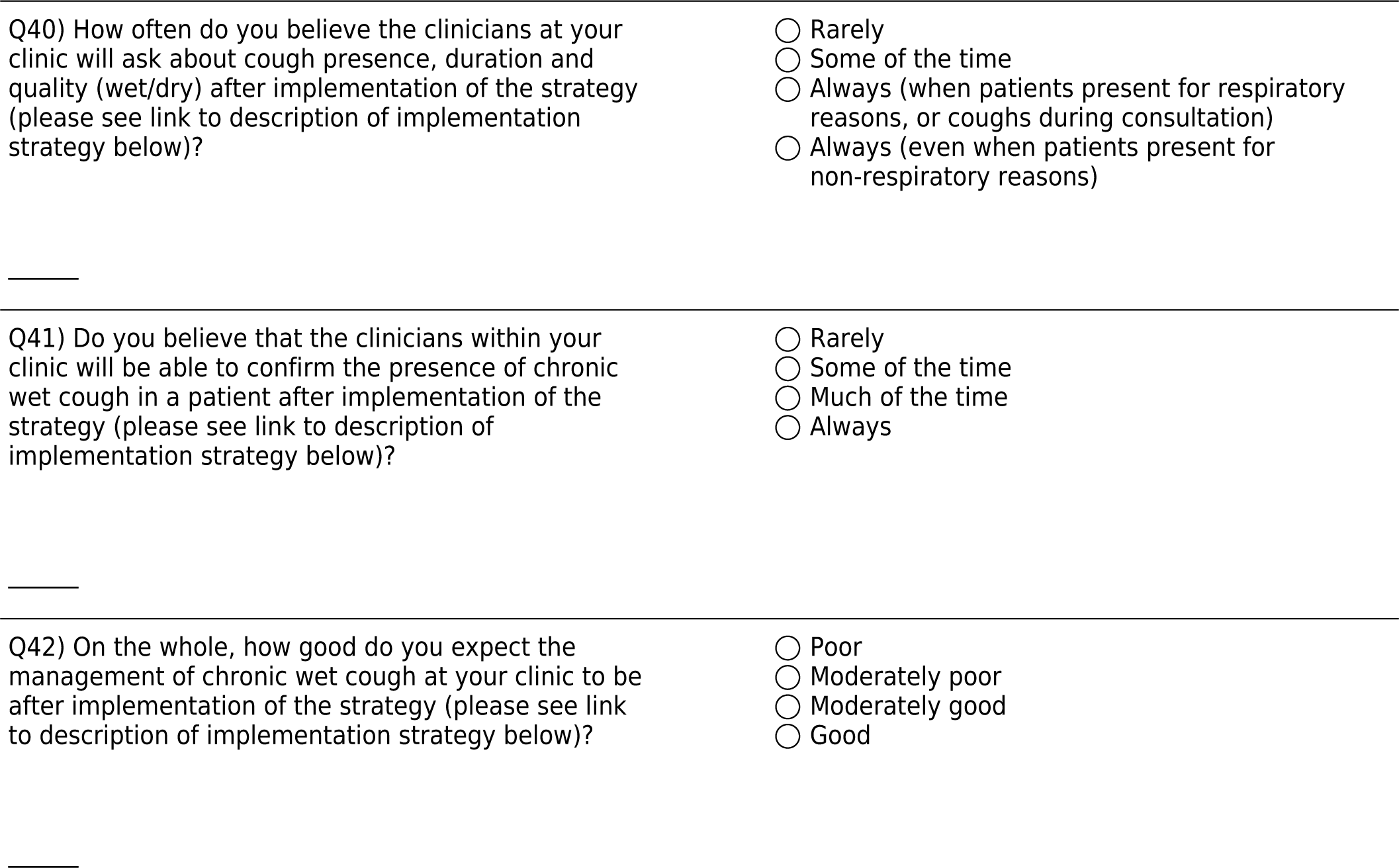

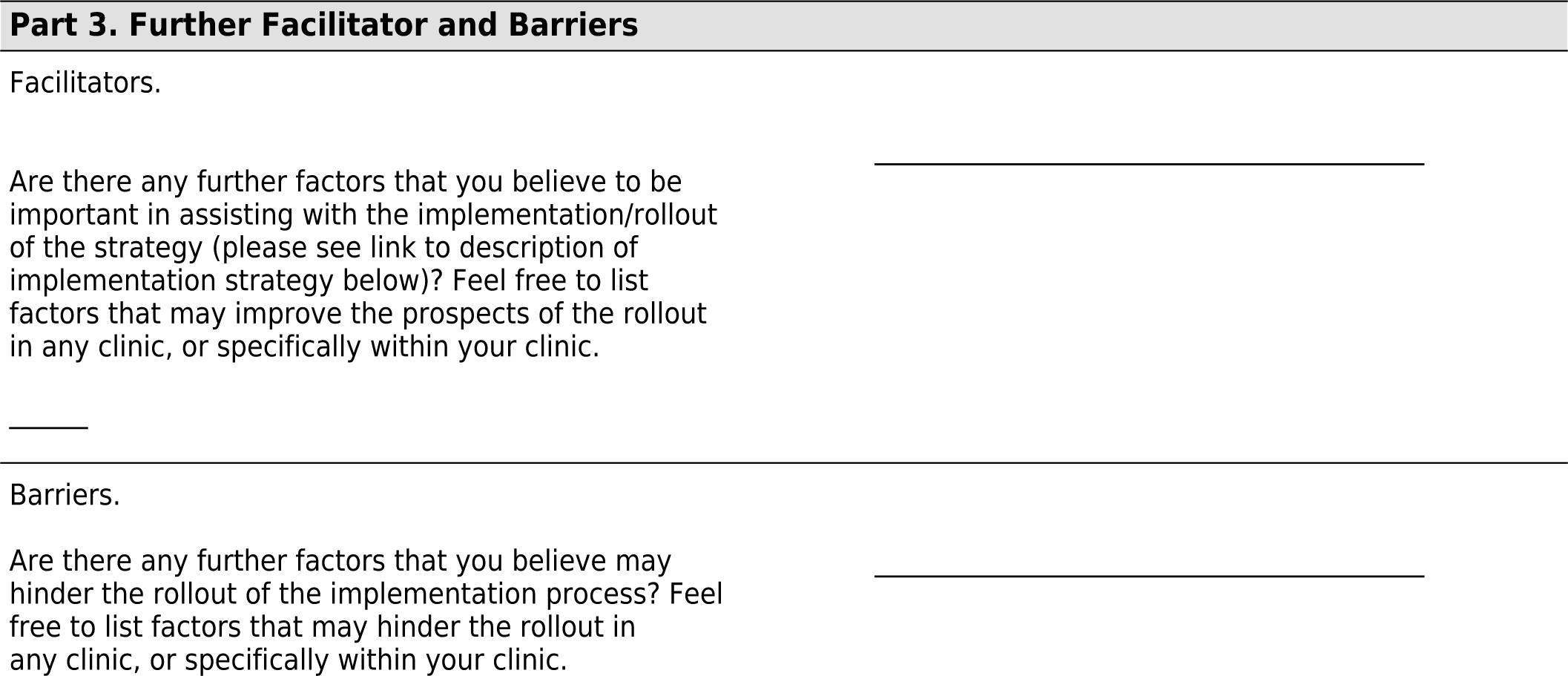

## Notes

### Competing Interest Statement

The authors have declared no competing interest.

### Funding Statement

The project was funded through a National Health and Medical Research Council Partnership Grant GTN1170735 and a Medical Research Future Fund Investigator Grant MRF1193796.

### Author Declarations

Ethics approval for our study was obtained from the Child and Adolescent Health Service Ethics Committee (RGS 4136), Western Australian Aboriginal Health Ethics Committee (HREC 774), Human Research Ethics Committee of the Northern Territory Department of Health and Menzies School of Health Research (HREC 2021-3954), and reciprocal ethics was given by the University of Queensland Human Research Ethics Committee (2020002563).

## References

Ashcraft, L., Goodrich, D., Hero, J., Phares, A., Bachrach, R., Quinn, D., Qureshi, N., Ernecoff, N., Scheunemann, L., Rogal, S., and Chinman, M. (2024). A systematic review of experimentally tested implementation strategies across health and human service settings: evidence from 2010-2022. Implementation Science, 19(43).

Balas, E. A. and Boren, S. A. (2000). Managing clinical knowledge for health care improvement. Yearbook of medical informatics, 9(01):65–70.

Barwick, M. (2023). The Implementation Roadmap. The Hospital for Sick Children, Toronto, Canada. https://web.cvent.com/event/6615d5da-45dc-4287-b0b6-097aa17bc83a/summary. Accessed: 2020-09-14.

Chang, A. B., Bush, A., and Grimwood, K. (2018). Bronchiectasis in children: diagnosis and treatment. Lancet, 392(10150):866–879.

Damschroder, L. J., Reardon, C. M., Widerquist, M. A. O., and Lowery, J. (2022). The updated consolidated framework for implementation research based on user feedback. Implement Sci, 17(1):75

Damschroder, Laura J Reardon, Caitlin M Widerquist, Marilla A Opra Lowery, Julie eng Research Support, U.S. Gov’t, Non-P.H.S. Review England 2022/10/31 Implement Sci. 2022 Oct 29;17(1):75. doi: 10.1186/s13012-022-01245-0.

Dempster, A. P., Laird, N. M., and Rubin, D. B. (1977). Maximum likelihood from incomplete data via the EM algorithm. Journal of the Royal Statistical Society: Series B (Methodological), 39(1):1–22.

D’Sylva, P., Walker, R., Lane, M., Chang, A. B., and Schultz, A. (2019). Chronic wet cough in aboriginal children: It’s not just a cough. Journal of paediatrics and child health, 55(7):833–843.

Fernandez, M. E., Ten Hoor, G. A., van Lieshout, S., Rodriguez, S. A., Beidas, R. S., Parcel, G., Ruiter, R. A. C., Markham, C. M., and Kok, G. (2019). Implementation mapping: Using intervention mapping to develop implementation strategies. Front. Public Health, 7:158.

Friedman, N. and Goldszmidt, M. (1998). Learning Bayesian Networks with Local Structure. In Jordan, M. I., editor, Learning in Graphical Models, pages 421–459. Springer Netherlands, Dordrecht.

Howard, R. A. and Matheson, J. E. (1981). Influence diagrams. In Howard, R. A. and Matheson, J. E., editors, Readings on the Principles and Applications of Decision Analysis, volume 2, pages 721–762. Strategic Decisions Group.

Institute of Medicine (2001). Crossing the Quality Chasm: A New Health System for the 21st Century. The National Academies Press, Washington, DC.

Kazdin, A. E. (2007). Mediators and mechanisms of change in psychotherapy research. Annu. Rev. Clin. Psychol., 3(1):1–27.

Korb, K. B. and Nicholson, A. E. (2010). Bayesian Artificial Intelligence. CRC Press, Inc., 2 edition.

Laird, P., Ball, N., Brahim, S., Brown, H., Chang, A. B., Cooper, M., Cox, D., Cox, D., Crute, S., Foong, R. E., Isaacs, J., Jacky, J., Lau, G., McKinnon, E., Scanlon, A., Smith, E. F., Thomason, S., Walker, R., and Schultz, A. (2022a). Prevalence of chronic respiratory diseases in aboriginal children: A whole population study. Pediatr Pulmonol, 57(12):3136–3144.

Laird, P., Walker, R., Lane, M., Chang, A. B., and Schultz, A. (2019). We won’t find what we don’t look for: Identifying barriers and enablers of chronic wet cough in aboriginal children. Respirology.

Laird, P., Walker, R., Lane, M., Totterdell, J., Chang, A. B., and Schultz, A. (2021). Recognition and management of protracted bacterial bronchitis in australian aboriginal children: A knowledge translation approach. Chest, 159(1):249–258.

Laird, P. J., Walker, R., McCallum, G., Toombs, M., Barwick, M., Morris, P., Aitken, R., Cooper, M., Norman, R., Patel, B., Lau, G., Chang, A. B., and Schultz, A. (2022b). Change in health outcomes for first nations children with chronic wet cough: rationale and study protocol for a multi-centre implementation science study. BMC Pulm Med, 22(1):492.

Lewis, C. C., Boyd, M. R., Walsh-Bailey, C., Lyon, A. R., Beidas, R., Mittman, B., Aarons, G. A., Weiner, B. J., and Chambers, D. A. (2020). A systematic review of empirical studies examining mechanisms of implementation in health. Implement. Sci., 15(1):21.

Linstone, H. A. and Turoff, M. (1975). Delphi Method: Techniques and Applications. Reading, Mass. Addison-Wesley Pub. Co., Advanced Book Program.

Loi, M. L. A. (2025). Diversity of aboriginal and torres strait islander culture. Accessed: 2025–02-19.

Mascaro, S. and Woodberry, O. (2022). A flexible method for parameterizing ranked nodes in Bayesian networks using Beta distributions. Risk Analysis, 42(6):1179–1195.

Mascaro, S., Woodberry, O., Wu, Y., and Nicholson, A. E. (2024a). The practice of qualitative parameterisation in the development of Bayesian networks.

Mascaro, S., Wu, Y., Pearson, R., Woodberry, O., Ramsay, J., Snelling, T., and Nicholson, A. E. (2024b). Causal knowledge engineering: A case study from COVID-19. arXiv:2403.14100 [cs].

Newcombe, P. A., Sheffield, J. K., and Chang, A. B. (2013). Parent cough-specific quality of life: development and validation of a short form. J Allergy Clin Immunol, 131(4):1069–74.

Newcombe, Peter A Sheffield, Jeanie K Chang, Anne B eng Research Support, Non-U.S. Gov’t 2012/11/14 J Allergy Clin Immunol. 2013 Apr;131(4):1069-74. doi: 10.1016/j.jaci.2012.10.004. Epub 2012 Nov 10.

Pearl, J. (1988). *Probabilistic reasoning in intelligent systems: networks of plausible inference*. Morgan Kaufmann, San Francisco.

Perry, C., Damschroder, L., Hemler, J., Woodson, T., Ono, S., and Cohen, D. (2019). Specifying and comparing implementation strategies across seven large implementation interventions: a practical application of theory. 14(1).

Proctor, E., Silmere, H., Raghavan, R., Hovmand, P., Aarons, G., Bunger, A., Griffey, R., and Hensley, M. (2011). Outcomes for implementation research: conceptual distinctions, measurement challenges, and research agenda. Adm. Policy Ment. Health, 38(2):65–76.

Smith, J. D., Li, D. H., and Rafferty, M. R. (2020). The implementation research logic model: a method for planning, executing, reporting, and synthesizing implementation projects. Implement. Sci., 15(1):84.

Spiegelhalter, D. J. and Lauritzen, S. L. (1990). Sequential updating of conditional probabilities on directed graphical structures. Networks. An International Journal, 20(5):579–605.

Vejnoska, S. F., Mettert, K., and Lewis, C. C. (2022). Mechanisms of implementation: An appraisal of causal pathways presented at the 5th biennial society for implementation research collaboration (SIRC) conference. Implement. Res. Pract., 3:26334895221086271.

Williams, N. J. (2016). Multilevel mechanisms of implementation strategies in mental health: Integrating theory, research, and practice. Adm. Policy Ment. Health, 43(5):783–798.

Zhang, N. L. and Yan, L. (1998). Independence of causal influence and clique tree propagation. International Journal of Approximate Reasoning, 19(3):335–349.

